# Breaking the Norm: Population-Scale Deviations of Brain Structure in Depression and Anxiety

**DOI:** 10.1101/2025.09.26.25336528

**Authors:** Julius Wiegert, Sebastián Marty-Lombardi, Jailan Oweda, Esra Lenz, Peter Ahnert, Klaus Berger, Hermann Brenner, Josef Frank, Hans J. Grabe, Karin Halina Greiser, Johanna Klinger-König, André Karch, Michael Leitzmann, Claudia Meinke-Franze, Rafael Mikolajczyk, Frauke Nees, Thoralf Niendorf, Oliver Sander, Carsten Oliver Schmidt, Steffi G. Riedel-Heller, Kerstin Ritter, Annette Peters, Tobias Pischon, Stephanie Witt, Johannes Nitsche, Joonas Naamanka, Sebastian Volkmer, Antonia Mai, Amrou Abas, Xiuzhi Li, Andreas Meyer-Lindenberg, Tobias Gradinger, Fabian Streit, Urs Braun, Emanuel Schwarz

## Abstract

Structural brain alterations associated with depression and anxiety are subtle, heterogeneous, and difficult to characterize. We applied autoencoder-based normative modeling to contrastively learned structural MRI representations from two large population-based cohorts (German National Cohort, *N* ≈ 29,000; UK Biobank, *N* ≈ 25,000) to quantify individual deviations from normative brain structure across symptom dimensions of depression, anxiety, and, for contextualization, alcohol use.

Deviation magnitude increased with symptom severity for depressive and anxiety symptoms and was most pronounced in individuals with high alcohol use. Directional analyses revealed shared deviation patterns for depression and anxiety that were largely distinct from alcohol-related deviations, and these patterns generalized across cohorts. These affective-symptom-related patterns implicated distributed regional brain-structural variation. Individual deviation profiles improved classification of symptomatic status beyond demographic covariates, with gains concentrated at higher symptom severity.

Together, these findings indicate that affective symptoms are associated with reproducible, dimensional patterns of regional brain-structural deviation that extend beyond normative population variability, supporting transdiagnostic models of internalizing psychopathology.

## Introduction

Major depressive disorder (MDD) and generalized anxiety disorder (GAD) are among the most prevalent psychiatric conditions worldwide and contribute substantially to the global burden of disease [1, 2, 3]. Both, MDD and GAD are characterized by disturbances across mood, motivation, cognition, and sleep, and are associated with substantial functional impairment and increased mortality risk [4, 5, 6]. Despite distinct diagnostic criteria, both disorders affect overlapping psychological domains and co-occur frequently.

Although MDD and GAD are defined as distinct diagnostic entities, they show substantial overlap in symptom domains, treatment response, and biological risk factors. Comorbidity rates reach up to 26% [7], and genetic correlations are high [8]. Evidence from neuroimaging further suggests shared neural substrates [9], complicating attempts to disentangle disorder-specific mechanisms. Both disorders exhibit pronounced clinical heterogeneity, including variation in symptom profiles, age of onset, and disease course [10, 11].

This clinical heterogeneity is mirrored on the biological level, particularly in attempts to predict MDD using brain imaging. A recent ENIGMA benchmark demonstrated that predicting MDD from structural MRI (sMRI) remains highly challenging [12]. Conventional supervised learning approaches implicitly assume biological similarity within diagnostic categories and separability between diagnoses, while also failing to exploit the large availability of healthy imaging data in population-based cohorts.

Normative modeling provides a complementary framework for studying psychiatric and neurodegenerative disorders by characterizing individual variation relative to a reference model learned from healthy populations [13, 14]. This approach enables the quantification of subject-specific deviations and has been applied across a wide range of conditions, including autism, dementia, and mood disorders [15, 16, 17, 18]. However, depending on the underlying feature representation, normative models may emphasize localized or summary-level deviations (e.g., voxel- or region-of-interest–based), which can limit their ability to capture highly distributed patterns of structural variation.

In this work, we pursued three aims: first, to characterize distributed brain-structural variation associated with depressive and anxiety symptoms; second, to quantify individual deviation magnitude relative to normative variability; and third, to assess the directional organization of these deviations using alcohol-related symptoms as a comparator, given their well-documented structural brain alterations [19, 20]. Together, these aims target both the magnitude and organization of brain-structural deviations while addressing transdiagnostic and individual heterogeneity.

To address these aims, we adopted a two-stage modeling strategy. First, we learned data-driven structural MRI representations from minimally processed sMRI data using contrastive representation learning. Recent contrastive approaches enable end-to-end feature learning without reliance on predefined regional summaries [21], and by comparing brain images at the population level, they may encourage representations that are sensitive to inter-individual differences expressed across distributed brain regions. Second, we applied autoencoder-based deep normative modeling to these contrastively learned representations, modeling the distribution of variation observed in healthy individuals and quantifying subject-specific deviations in latent space [22, 23]. This separation of representation learning and normative modeling allows the normative model to operate on a compact, data-driven embedding of distributed brain-structural differences.

Depressive and anxiety symptoms are present in the general population [24]. Thus we leveraged population-scale cohorts from the German National Cohort (NAKO) and the UK Biobank (UKB) [25, 26]. sMRI is available at scale in both cohorts, exhibits high test–retest reliability [27], and provides a stable readout of cumulative neurobiological variation well suited for large-scale normative modeling. Moreover, large meta-analyses demonstrate that gray matter volume (GMV) alterations in depression and anxiety disorders are spatially distributed rather than regionally focal [28, 29]. The NAKO cohort served as the discovery dataset, with external validation performed in the UK Biobank.

## Materials and Methods

This study was approved by the Ethics Committee II of the University of Heidelberg Medical Faculty Mannheim (protocol no. 2025-827). NAKO and UKB data collection were approved by the relevant local ethics committees in Germany and the United Kingdom and were conducted in accordance with the Declaration of Helsinki. All participants provided written informed consent. Unless stated otherwise, all model training was performed in the NAKO cohort; UK Biobank data were used exclusively for evaluation.

### Group Definitions

Current depressive symptoms were assessed using the depression scale of the Patient Health Questionnaire-9 (PHQ-9) [30], anxiety symptoms using the anxiety scale of the Generalized Anxiety Disorder-7 (GAD-7) [31], and alcohol use using the consumption scale of the Alcohol Use Disorders Identification Test–Consumption (AUDIT-C) [32]. These instruments were selected because they provide ordinal severity measures available in both cohorts.

Participants were stratified into symptom severity groups based on established cutoff scores to define interpretable severity strata: PHQ-9 none/minimal (0–4), mild (5–9), moderate (10–14), moderately severe (15–19), and severe (20–27) [30]; GAD-7 minimal (0–4), mild (5–9), moderate (10–14), and severe (15–21) [31]; and AUDIT-C low (0–4), increasing (5–7), higher risk (8–10), and possible dependence (11–12) [32]. To ensure sufficient statistical power while retaining resolution at higher symptom levels, individuals with AUDIT-C scores above 7 were further split into groups with scores of 8, 9, and *≥*10.

Healthy controls (HCs) were defined using harmonized thresholds across instruments (PHQ-9 *<* 5, GAD-7 *<* 5, AUDIT-C *<* 8). To minimize psychiatric contamination, individuals with lifetime or self-reported diagnoses of mood, anxiety, or alcohol use disorders were excluded using cohort-specific diagnostic information.

### Cohorts - German National Cohort (NAKO)

The NAKO is a population-based study designed to investigate the causes and early stages of common chronic diseases [25]. The study enrolled 205,000 participants (aged 20–69 years) from 18 centers across Germany, of whom 30,861 underwent MRI at five centers with dedicated facilities [33]. The baseline assessment, conducted between 2014 and 2019, comprised a comprehensive list of examinations, standardized face-to-face medical interviews and touchscreen-based self-report questionnaires, as well as biomaterial collection.

We performed an initial data filtering step by excluding samples with missing questionnaire data (PHQ-9, GAD-7, or AUDIT-C), diagnostic data (MINI or self-reported diagnosis), or confounder data (age, sex, or scan site). Participants with a history of stroke (N = 301) or Parkinson’s disease (N = 23) were also excluded to reduce potential bias from neurodegenerative conditions. In total, 1,570 samples were excluded due to missing questionnaire or diagnostic data. The final group distributions and demographics used in subsequent analyses are summarized in Table 1.

**Table 1:** Demographics in the NAKO and UKB cohort. Abbreviations: HC, healthy controls; AUDIT-C, Alcohol Use Disorders Identification Test–Consumption; PHQ-9, Patient Health Questionnaire-9; MDD, major depressive disorder; MINI, MINI International Neuropsychiatric Interview; GAD-7, Generalized Anxiety Disorder-7; GAD, generalized anxiety disorder; ANX/PD, anxiety or panic disorder; ICD-10, International Classification of Diseases, 10th Revision.

| Group | Criterion | Count | Age $\mu(\pm\sigma)$ | Females (%) |
| --- | --- | --- | --- | --- |
| **NAKO cohort** |  |  |  |  |
| HC | – | 15 941 | 48 ($\pm 12$) | 39 |
| AUDIT-C (8) | AUDIT-C sum = 8 | 633 | 49 ($\pm 13$) | 15 |
| AUDIT-C (9) | AUDIT-C sum = 9 | 249 | 49 ($\pm 13$) | 10 |
| AUDIT-C $\geq 10$ | AUDIT-C sum $\geq 10$ | 111 | 51 ($\pm 13$) | 11 |
| MDD (mild) | PHQ-9 sum $\in [5, 9]$ | 6 829 | 51 ($\pm 11$) | 53 |
| MDD (moderate) | PHQ-9 sum $\in [10, 14]$ | 1 464 | 46 ($\pm 12$) | 56 |
| MDD (moderately severe) | PHQ-9 sum $\in [15, 19]$ | 423 | 45 ($\pm 12$) | 55 |
| MDD (severe) | PHQ-9 sum $\geq 20$ | 181 | 47 ($\pm 12$) | 55 |
| MDD (MINI) | MINI Diagnosis MDD | 4 752 | 59 ($\pm 11$) | 54 |
| MDD (Diagnosis) | Doctor’s Diagnosis | 3 636 | 50 ($\pm 11$) | 60 |
| GAD (mild) | GAD-7 sum $\in [5, 9]$ | 5 566 | 47 ($\pm 12$) | 54 |
| GAD (moderate) | GAD-7 sum $\in [10, 14]$ | 1 058 | 46 ($\pm 12$) | 56 |
| GAD (severe) | GAD-7 sum $\geq 15$ | 320 | 46 ($\pm 12$) | 59 |
| ANX/PD (Diagnosis) | Doctor’s Diagnosis | 1 716 | 49 ($\pm 12$) | 59 |
| **UKB cohort** |  |  |  |  |
| HC | – | 13 582 | 65 ($\pm 7$) | 53 |
| AUDIT-C (8) | AUDIT-C sum = 8 | 845 | 63 ($\pm 7$) | 32 |
| AUDIT-C (9) | AUDIT-C sum = 9 | 611 | 62 ($\pm 7$) | 25 |
| AUDIT-C $\geq 10$ | AUDIT-C sum $\geq 10$ | 631 | 63 ($\pm 7$) | 22 |
| AUD | ICD-10 | 192 | 64 ($\pm 8$) | 25 |
| MDD (mild) | PHQ-9 sum $\in [5, 9]$ | 3 357 | 61 ($\pm 7$) | 63 |
| MDD (moderate) | PHQ-9 sum $\in [10, 14]$ | 770 | 61 ($\pm 8$) | 62 |
| MDD (moderately severe) | PHQ-9 sum $\in [15, 19]$ | 267 | 60 ($\pm 7$) | 66 |
| MDD (severe) | PHQ-9 sum $\geq 20$ | 132 | 58 ($\pm 6$) | 70 |
| MDD (ICD-10) | ICD-10 | 754 | 62 ($\pm 8$) | 65 |
| MDD (Diagnosis) | Doctor’s Diagnosis | 3 918 | 63 ($\pm 7$) | 66 |
| GAD (mild) | GAD-7 sum $\in [5, 9]$ | 2 927 | 61 ($\pm 8$) | 66 |
| GAD (moderate) | GAD-7 sum $\in [10, 14]$ | 559 | 61 ($\pm 8$) | 65 |
| GAD (severe) | GAD-7 sum $\in [15, 21]$ | 276 | 59 ($\pm 7$) | 67 |
| GAD (ICD-10) | ICD-10 | 19 | 63 ($\pm 7$) | 66 |
| ANX (ICD-10) | ICD-10 | 654 | 63 ($\pm 7$) | 65 |
| ANX (Diagnosis) | Doctor’s Diagnosis | 2 646 | 63 ($\pm 7$) | 63 |

### Cohorts - UK Biobank (UKB)

The UKB is a population-based cohort of approximately 500,000 participants aged 40–69 years at recruitment, with sMRI data available for 49,279 individuals [26]. Individuals with neurodegenerative diseases (N = 2,589; ICD-10 codes) were excluded. After further excluding participants with missing questionnaire data (PHQ-9, GAD-7, or AUDIT-C) or missing confounder information (age, sex, or total intracranial volume (TIV)), 24,838 individuals remained (Table 1).

Because PHQ-9 and GAD-7 scores were available in UKB, the same HC criteria were applied as in NAKO. However, questionnaire assessments in UKB were completed independently of the MRI scan date [34], resulting in substantially larger temporal offsets between imaging and symptom measures (median absolute deviation: 742 days in UKB vs. 15 days in NAKO). This temporal mismatch represents an inherent limitation. In addition to questionnaire-based definitions, ICD- 10 diagnoses and self-reported diagnoses for MDD, GAD, ANX, and AUD were included in UKB analyses.

### Magnetic Resonance Imaging

Structural T1-weighted MRI data were analyzed. In NAKO, images were acquired using 3D MPRAGE (TR = 2300 ms, TE = 2.98 ms, TI = 900 ms) on a 3T Siemens Skyra scanner with 1 mm isotropic resolution. Images were processed using CAT12 (v12.9), including bias correction, segmentation, normalization to MNI152NLin2009cAsym space, modulation, and resampling. For computational tractability while preserving macro-anatomical structure relevant for whole-brain representation learning, GMV images were downsampled to a standardized 80 × 80 × 80 voxel grid using spline interpolation, a resolution successfully used in 3D CNN-based brain MRI studies [35, 36]. Voxel intensities were subsequently standardized.

In UKB, T1-weighted images were acquired using 3D MPRAGE (TR = 2000 ms, TE = 2.01–2.03 ms, TI = 880 ms) on a 3T Siemens scanner. Images were preprocessed using FSL (v5.0.10) FAST and registered to the same MNI template using FLIRT. Identical downsampling and standardization procedures were applied to ensure comparability across cohorts.

Site- and scanner-related variation was addressed using linear residualization and additionally, where appropriate, by including the corresponding variables as covariates in downstream analyses.

### Overview of the Analytical Pipeline

In brief, the analytical pipeline consisted of three steps. First, self-supervised contrastive learning was used to transform each sMRI scan into a compact representation of distributed inter-individual brain-structural variation. Second, an autoencoder trained exclusively on HCs was used to model the normative range of these representations; deviations were defined as the difference between observed and reconstructed embeddings. Third, these deviation profiles were analyzed with respect to their magnitude, direction, regional correlates, and ability to classify symptom-defined groups. Thus, the analyses quantify departures from a learned multivariate structural reference space rather than focal voxelwise abnormalities or diagnostic markers. Details are provided below and in the Supplementary Material.

### Contrastive Feature Extraction using Momentum Contrast

A schematic overview of the representation learning and normative modeling pipeline is provided in Supplementary Figure 1.

To derive compact, multivariate representations of inter-individual variation in brain structure, we applied momentum contrast (MoCo) for self-supervised feature extraction on GMV images [37], using a three-dimensional convolutional neural network (3D CNN) encoder. MoCo was trained on the full NAKO MRI dataset, including HCs and symptomatic participants. As training was fully unsupervised and did not use diagnostic information, this procedure does not introduce label leakage and follows standard practice in representation learning. MoCo was trained to produce 256-dimensional embeddings, a commonly used embedding size in prior contrastive learning work that balances representational capacity with downstream model complexity [38, 39, 40].

All deep learning models were implemented in PyTorch (version 2.2.0) [41].

### Normative Modeling using Autoencoder

MoCo embeddings were linearly residualized with respect to covariates age, age squared, sex, scanning site, and TIV to reduce variance attributable to non-pathological sources, and subsequently min–max scaled to ensure numerical stability.

The residualized embeddings were input to a fully connected autoencoder trained exclusively on HCs from the NAKO cohort. HC data were split into training, validation, and test sets (70/15/15%), ensuring model selection and generalization assessment without exposure to symptomatic subjects. The autoencoder mapped the 256-dimensional input to a 50-dimensional bottleneck and back to the original space, and reconstruction error-derived deviations were interpreted relative to the learned normative manifold rather than as probabilistic scores. Architectural details, bottleneck selection, optimization settings, and training/validation diagnostics are provided in Supplementary Sections 2.1–2.3 and Supplementary Tables 8–9.

Normative deviations were quantified using reconstruction errors from the HC-trained autoen-coder. Because the autoencoder partially reintroduced confounding structure, deviation vectors were subjected to a second linear residualization using the same covariates. Prior to residualization, deviations showed widespread associations with age, age squared, sex, and age *×* sex; after residualization, only a single weak association with sex remained.

### Shift Analysis of Mahalanobis Distances

To quantify how symptomatic groups deviate from normative variability at the group level, we performed a shift analysis comparing the full distribution of deviations rather than relying on summary statistics such as central tendencies.

For each cohort (NAKO and UKB), Mahalanobis distances in the 256-dimensional deviation space were computed relative to a cohort-specific HC reference distribution. For an individual deviation vector *x*, the Mahalanobis distance was defined as

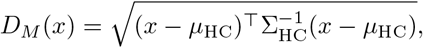

where *µ*_HC_ and Σ_HC_ denote the mean vector and covariance matrix estimated from the HC reference sample, respectively [42]. This yielded a covariance-adjusted summary of how strongly each individual’s deviation profile departed from the HC reference distribution. The mean vector and covariance matrix were estimated from a random 90% subset of HCs, and distances were subsequently computed for symptomatic participants and the remaining 10% HC holdout set.

Cohort-specific HC reference distributions were used because Mahalanobis distances depend on the mean and covariance structure of the reference population. Accordingly, absolute deviation magnitudes are not directly comparable across cohorts. Stability of reference parameters was assessed by bootstrap resampling (Supplementary Section 2.5).

Distributional shift was quantified as the exponentially weighted area in which the Mahalanobis distance distribution of a symptomatic group exceeded that of HCs. This weighting emphasizes larger deviations, yielding a scalar metric between 0 and 1 that summarizes the extent to which symptomatic variability exceeds normative heterogeneity (details in Supplementary Section 2.4). As this metric is designed to capture distribution-level differences, it is intended for group-level comparison.

Uncertainty in shift estimates was quantified using nonparametric bootstrap resampling (1,000 iterations) of subjects within each group, from which 95% confidence intervals were derived.

### Significance Testing of Mahalanobis Distances

To assess covariate-adjusted group differences in deviation magnitude, we performed individual-level inference using linear regression models with heteroskedasticity-consistent (HC3) standard errors. For each symptomatic group, log-transformed Mahalanobis distances were regressed on binary group membership (symptomatic vs. HC).

Regression-based inference was used to allow covariate adjustment. Log-transformation and robust standard errors were used to mitigate non-normality and the influence of extreme values.

Statistical significance of group membership was assessed using type III ANOVA on the fitted models, performed separately for each symptomatic group. Resulting *p*-values reflect covariate-adjusted differences in individual deviation magnitude between symptomatic participants and HCs and complement the distributional shift analysis. To control for multiple testing across diagnostic groups, *p*-values were corrected using the Benjamini–Hochberg false discovery rate (FDR) procedure. Unless stated otherwise, significance was assessed at an FDR threshold of *α* = 0.05.

### Directional Analysis

In addition to the scalar shift analysis, we examined the geometry of normative deviations after projection into a low-dimensional space for visualization. Deviations were defined as the difference between each subject’s original and reconstructed feature representations in the deconfounded 256-dimensional deviation space.

For visualization, subject-level deviation vectors were standardized and projected into two dimensions using principal component analysis (PCA). The first two components explained 21% of the variance. Subjects were then grouped according to symptom domain (PHQ-9, GAD-7, AUDIT-C), severity level, and comorbidity status.

Robust linear models correcting for covariates were fitted in symptomatic subjects only, relating each principal deviation score (PC1, PC2) to symptom severity (PHQ-9 or GAD-7) in separate models, with HC3 standard errors. Multiple testing was controlled using Benjamini–Hochberg FDR correction. Effect sizes were summarized using standardized regression coefficients (*β*_std_).

Group-level deviation vectors were estimated as geometric medians. Stability was assessed using 1,000 bootstrap resamples, from which 1-SD uncertainty ellipses were derived.

All group-level deviation vectors and bootstrap ellipses were centered relative to the HC centroid. This analysis is intended as a qualitative assessment of relative deviation directions and overlap between groups in the projected space, rather than a formal inferential test in the full high-dimensional representation.

### Explaining Normative Deviations

To relate the learned latent deviation dimensions to interpretable neuroanatomical features, we performed a post hoc, heuristic analysis linking deviations to regional gray matter measures. Direct gradient-based attribution methods (e.g., saliency maps) are not applicable in this setting because multiple deconfounding and residualization steps break the differentiable mapping between voxel-level inputs and final deviation estimates. Moreover, the normative model is spatially unconstrained and operates on distributed whole-brain representations rather than predefined regions. Accordingly, the following analysis is exploratory in nature and intended for hypothesis generation rather than definitive anatomical localization.

The analysis comprised three steps. First, deviation dimensions were screened for associations with symptom severity. Second, relationships between regional GMV measures and latent embedding dimensions were quantified. Third, symptom-associated deviation dimensions were aggregated to derive regional relevance maps.

In the first step, questionnaire sum scores (PHQ-9, GAD-7, AUDIT-C) were correlated with each of the 256 deviation dimensions using Spearman rank correlations. To account for multiple testing (3 *×* 256 tests), we applied the Storey–Tibshirani q-value procedure [43]. Statistical significance was defined as *q <* 0.05, corresponding to control of the positive FDR (pFDR).

In the second step, elastic net regression [44] was used to predict each covariate-adjusted MoCo embedding dimension from 99 covariate-adjusted FreeSurfer GMV features. Elastic net regularization was chosen for its robustness to multicollinearity and its ability to yield sparse and stable region–embedding associations.

In the third step, regional coefficients from the elastic net models were weighted by the corresponding significant dimension–symptom correlations and summed across all selected dimensions, yielding a single heuristic relevance score per brain region and symptom domain.

### Classification Analysis

To quantify the predictive value of the deviations beyond demographic effects, we performed classification analyses using center, age, age squared, sex, and the sex *×* age interaction as baseline covariates.

All classification analyses were performed exclusively in the UKB cohort, ensuring strict separation between representation learning and supervised evaluation.

We evaluated two model configurations: (i) confounders only, and (ii) normative deviation features plus confounders, where the latter included the 256-dimensional deviation vectors derived from the autoencoder.

Binary classifications (symptomatic group vs. HC) were performed using logistic regression with elastic net regularization. The elastic net mixing parameter (*𝓁*_1_ ratio) was tuned within each training fold using an inner stratified 80/20 split. To prevent data leakage, all features were standardized within each training fold using parameters estimated from the corresponding training data only.

Model performances were evaluated using stratified 10-fold cross-validation and summarized reporting the mean and standard deviation of the area under the ROC curve (AUC) and balanced accuracy (BACC), with a fixed decision threshold of 0.5.

## Results

### Shift Analysis

**Figure 1A** illustrates how deviations in symptomatic PHQ-9, GAD-7, and AUDIT-C groups diverge from normative variation defined by an independent HC holdout set (N=1,594). **Figure 1B** shows that the magnitude of deviation is associated with the likelihood of belonging to at least moderately symptomatic groups. We quantified the proportion of each group whose deviations exceeded the range observed among HCs (**Figure 2A**, henceforth referred to as “shift”).

**Figure 1:**
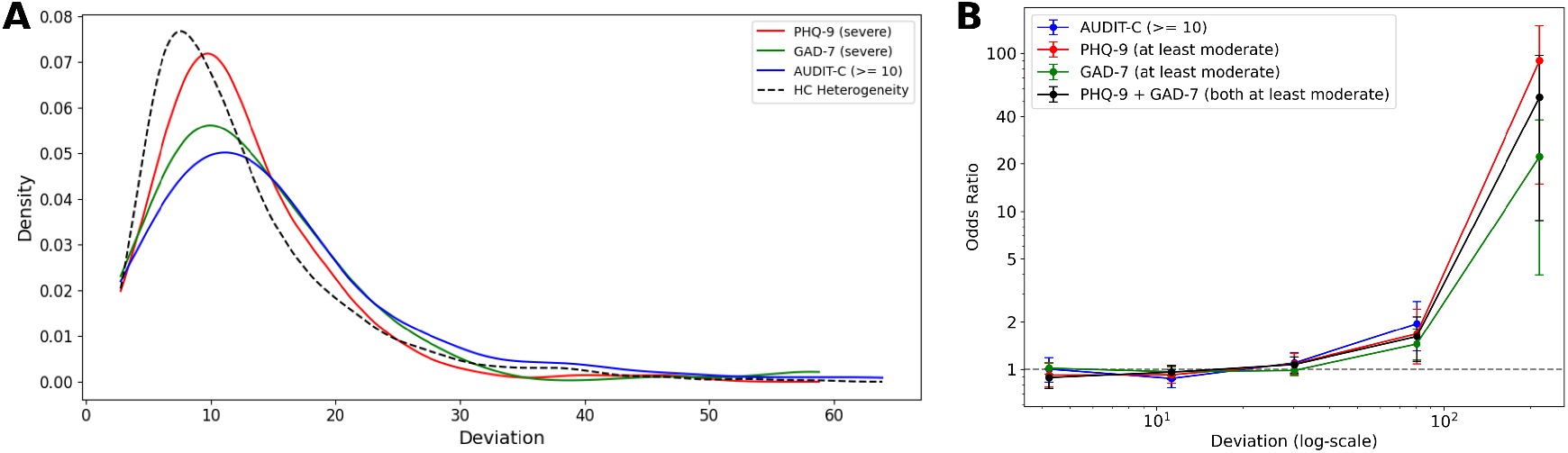
**A**: Density estimates of Mahalanobis distance (deviation) from the norm for three exemplary symptomatic groups. The dashed black line represents the natural heterogeneity among independent healthy controls (HCs). **B**: Relationship between deviation from the normative distribution and the odds ratio (OR) of having Patient Health Questionnaire-9 (PHQ-9) *≥*10, Generalized Anxiety Disorder-7 (GAD-7) *≥*10, and an Alcohol Use Disorders Identification Test–Consumption (AUDIT-C) *≥*10. Deviation scores were stratified into 5 bins with edges defined on a base-10 logarithmic scale, spanning the minimum to maximum observed distances. This ensured relatively finer resolution in the distribution tail where case enrichment was expected. A strong increase in OR with increasing deviation illustrates that individuals further from the norm are substantially more likely to be symptomatic. Error bars denote 95% confidence intervals obtained via 1000 bootstrap resamples. In the highest deviation bin, there was 1 HC, 0 AUDIT-C samples, 6 PHQ-9 samples, 7 GAD-7 samples, and 6 PHQ-9 + GAD-7 samples.

**Figure 2:**
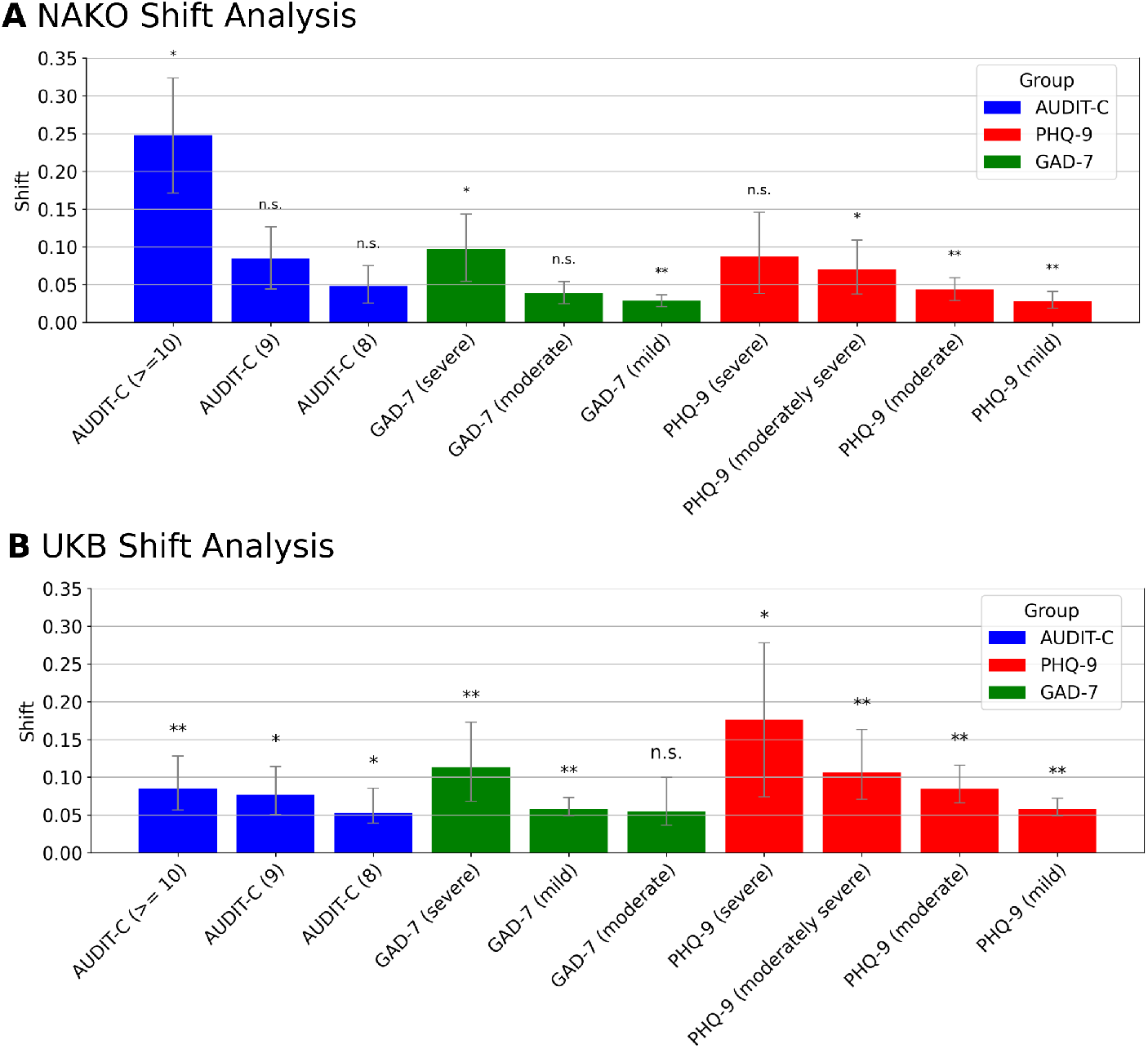
Fraction of symptomatic group deviations unexplained by healthy control variability (*shift*) for all symptomatic groups for NAKO cohort (German National Cohort) (**A**) and the UKB cohort (UK Biobank) (**B**). Because shifts were computed relative to cohort-specific HC reference distributions, values are not directly comparable as absolute deviation magnitudes across cohorts and should be interpreted within cohort. To assess statistical significance, underlying Mahalanobis distances were modeled as the dependent variable in a linear regression with group membership as the main predictor, adjusting for sex, age, age-squared, and sex–age interactions. Asterisks indicate the level of statistical significance after multiple testing correction using Benjamini-Hochberg correction [45]: * *p*_*F DR*_ *<* 0.05, ** *p*_*F DR*_ *<* 0.01. Error bars indicate 95% confidence intervals obtained from 1000 bootstrap samples. Symptomatic groups are defined by AUDIT-C (Alcohol Use Disorders Identification Test–Consumption), PHQ-9 (Patient Health Questionnaire-9), and GAD-7 (Generalized Anxiety Disorder-7) thresholds.

This analysis therefore asks whether symptomatic groups contain a larger fraction of individuals with deviation profiles outside the range typically observed among HCs.

At the symptomatic group level, participants with higher symptom severity levels for MDD and GAD (encoded on a scale from 1 [mild] to 4 [severe]) exhibited significantly larger shifts. Spearman rank correlations on the group level between symptom severity and shift revealed strong positive associations when jointly analyzing the PHQ-9 and GAD-7 groups (*ρ* = 0.92, p = 0.006), i.e., levels 1 to 4 for PHQ-9 and 2 to 4 for GAD-7 and their corresponding shift values.

The group with the highest shift was alcohol-related, with a shift of over 20% in the top AUDIT-C (*≥*10) group (**Figure 2 A**). Recent, severe depressive and anxiety symptoms exhibited larger deviations than lifetime diagnoses assessed via clinical interviews or self-report (Supplementary Figure 3). P-values for all groups are provided in Supplementary Tables 10 and 11. Sex-stratified analyses showed comparable trends (Supplementary Figure 4).

As shown in **Figure 2B**, deviation shifts in UKB followed a pattern that was qualitatively consistent with NAKO. Because Mahalanobis distances and shift estimates were defined relative to cohort-specific HC reference distributions, absolute deviation magnitudes are not directly comparable across cohorts. Cross-cohort comparisons are therefore interpreted descriptively, focusing on the relative ordering of symptom strata and the recurrence of severity-related patterns rather than on absolute differences in magnitude. Under this interpretation, the relative ordering of symptom strata by severity was broadly preserved across cohorts.

## Directional Analysis

To examine how psychiatric symptom profiles manifest in the space of normative brain deviations, we analyzed the geometry of group-level deviation vectors relative to the healthy reference. Participants were stratified by symptom domain (PHQ-9, GAD-7, AUDIT-C), severity level, and comorbidity status, allowing us to assess both the extent of heterogeneity within symptom groups and the presence of higher-level structural patterns across domains.

Group-representative deviation vectors were estimated as geometric medians and projected into two principal components for visualization (**Figure 3A**). Shaded ellipses reflect bootstrap uncertainty, highlighting substantial within-group heterogeneity.

**Figure 3:**
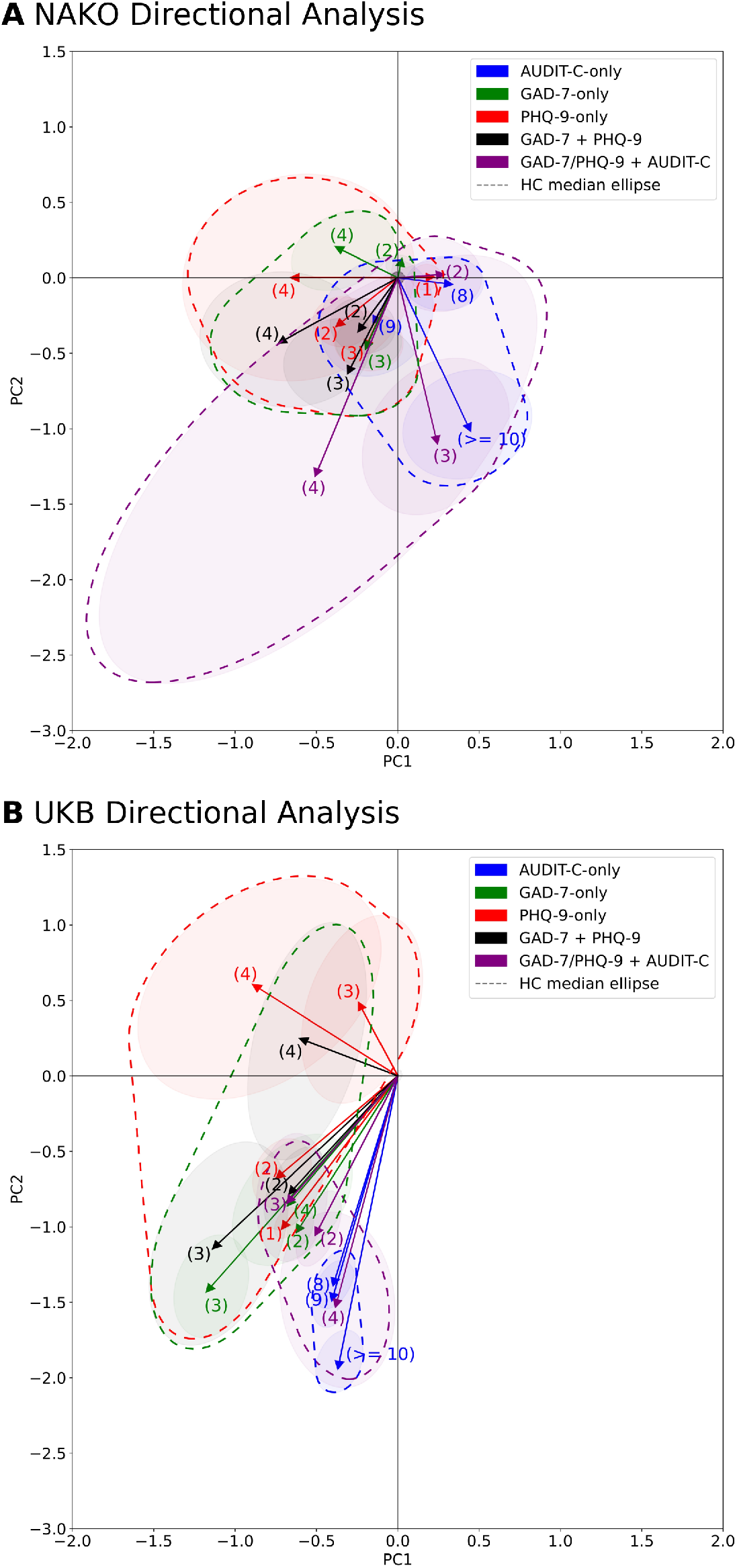
Directional analyses based on principal component analysis (two dimensions, 21% explained variance) in the NAKO cohort (German National Cohort) (**A**) and the UKB cohort (UK Biobank) (**B**). Shaded ellipses denote the ±1 SD contour of the bootstrap (1,000 resamples) distribution of the group-level geometric median vectors. The dark ellipse around the origin indicates the median deviation among healthy controls (HCs). Numbers indicate severity levels. “MDD/GAD + ALC” denotes individuals with either acute MDD or GAD symptoms and elevated alcohol use (AUDIT-C *≥*10). For PHQ-9, levels (1) to (4) correspond to mild, moderate, moderately severe, and severe symptoms, respectively. For GAD-7, levels (2) to (4) correspond to mild, moderate, and severe symptoms. The scattered area includes the ellipses of GAD, PHQ, GAD+PHQ groups (red), AUDIT-C groups (blue) and comorbid groups (purple). Because deviation vectors were centered and interpreted relative to cohort-specific HC references, apparent distances from the HC centroid should not be interpreted as directly comparable absolute deviation magnitudes across cohorts.

Despite substantial within-group heterogeneity, consistent meta-patterns emerged. Alcoholrelated groups deviated along a direction largely distinct from the mood–anxiety axis traced by PHQ-9 and GAD-7 groups, while increasing AUDIT-C thresholds showed progressively larger deviations. Depressive and anxiety symptom groups overlapped extensively, with higher symptom levels shifting further along a shared mood–anxiety-related direction. Individuals with concurrent PHQ-9/GAD-7 elevation occupied intermediate positions rather than forming a separable cluster, supporting a continuous spectrum of overlapping deviation patterns. Participants with elevated AUDIT-C and concurrent depressive or anxiety symptoms clustered between alcohol-related and mood–anxiety-associated directions, consistent with mixed or additive deviation profiles. Comparable patterns were observed for interview-based and self-reported diagnoses (Supplementary Figure 5).

A sensitivity analysis examining the stability of deviation directions under sex stratification is provided in Supplementary Figure 6.

In symptomatic subjects (*n* = 13,003), GAD-7 scores were associated with PC1 deviations (*q* = 0.003), whereas PHQ-9 scores were not (*q* = 0.077); conversely, both PHQ-9 (*q* = 0.004) and GAD-7 (*q* = 0.022) were associated with PC2. Effect sizes were uniformly small (standardized *β≈* 0.02–0.03). In the UKB external validation sample (symptomatic subjects-only; *n* = 9,010), PC1 was associated with both PHQ-9 (*β*_std_ = 0.030, *q* = 5.5 *×*10^−3^) and GAD-7 (*β*_std_ = *−* 0.033, *q* = 4.1 *×*10^−3^), whereas PC2 was associated only with GAD-7 (*β*_std_ = 0.040, *q* = 5.1*×* 10^−4^) and not with PHQ-9 (*q* = 0.054).

To assess whether the geometric relationships observed in NAKO generalize to an independent cohort, UKB deviation vectors were projected into the directional PCA deviation space (**Figure 3B**). Despite substantial within-group heterogeneity, several higher-level patterns are consistent across cohorts. Alcohol-related symptom groups again deviate along directions that are largely distinct from the mood–anxiety axis traced by PHQ-9 and GAD-7 groups. Mood and anxiety symptom strata occupy overlapping regions of the deviation space, and participants with concurrent mood–anxiety and alcohol-related symptom elevation fall in intermediate positions between the corresponding directions, which appears to be the case in our analysis.

At the same time, cohort-specific differences were visually evident. In UKB, symptom groups appeared more displaced from the HC centroid than in NAKO, including lower symptom-burden and alcohol-related strata. Because deviation vectors were centered and interpreted relative to cohort-specific HC references, this apparent displacement should be interpreted descriptively rather than as evidence for larger absolute deviation magnitudes in UKB. It may reflect differences in cohort composition, assessment context, population health, or residual cohort-specific effects, rather than a change in the relative geometry of deviation patterns.

Consistency of the directional patterns was further supported by analyses based on ICD-10 and self-reported diagnoses in the UKB (Supplementary Figure 5).

A sensitivity analysis stratifying UKB participants by MRI–questionnaire interval showed that group-level deviation directions were highly similar to the unfiltered estimates for both the shortest and longest interval quintiles when group sizes were sufficiently large, whereas estimates became unstable only under restrictive minimum-size thresholds with few retained groups (Supplementary Figure 7).

### Importance of Brain Regions

We first examined which latent deviation dimensions were associated with symptom severity. Of the 256 normative deviation dimensions, 14 were significantly associated with PHQ-9 scores (5.4%), 4 with GAD-7 scores (1.6%), and 35 with AUDIT-C scores (13.7%) after multiple-testing correction (q-value *<* 0.05).

This mapping should be interpreted as a heuristic summary of distributed associations between latent deviation dimensions and regional GMV features, rather than as evidence for focal anatomical effects. Elastic net models explained a moderate fraction of variance in the MoCo embedding dimensions (median *R*^2^ = 0.22, range = 0.10–0.40; Supplementary Table 14), further supporting cautious interpretation of the regional relevance maps.

Aggregating regional GMV associations via our heuristic mapping procedure yielded distributed regional relevance patterns rather than focal effects. Figure 4 shows the regions contributing most consistently to symptom-associated deviation dimensions. Across all three symptom domains, cerebellar regions emerged as the most prominent contributors. For AUDIT-C, these associations were predominantly negative, indicating that higher alcohol-related symptom severity was linked to deviation dimensions associated with lower cerebellar GMV. Additional negative contributions were observed in posterior cortical regions, including the cuneus (Supplementary Figures 11 and 12).

**Figure 4:**
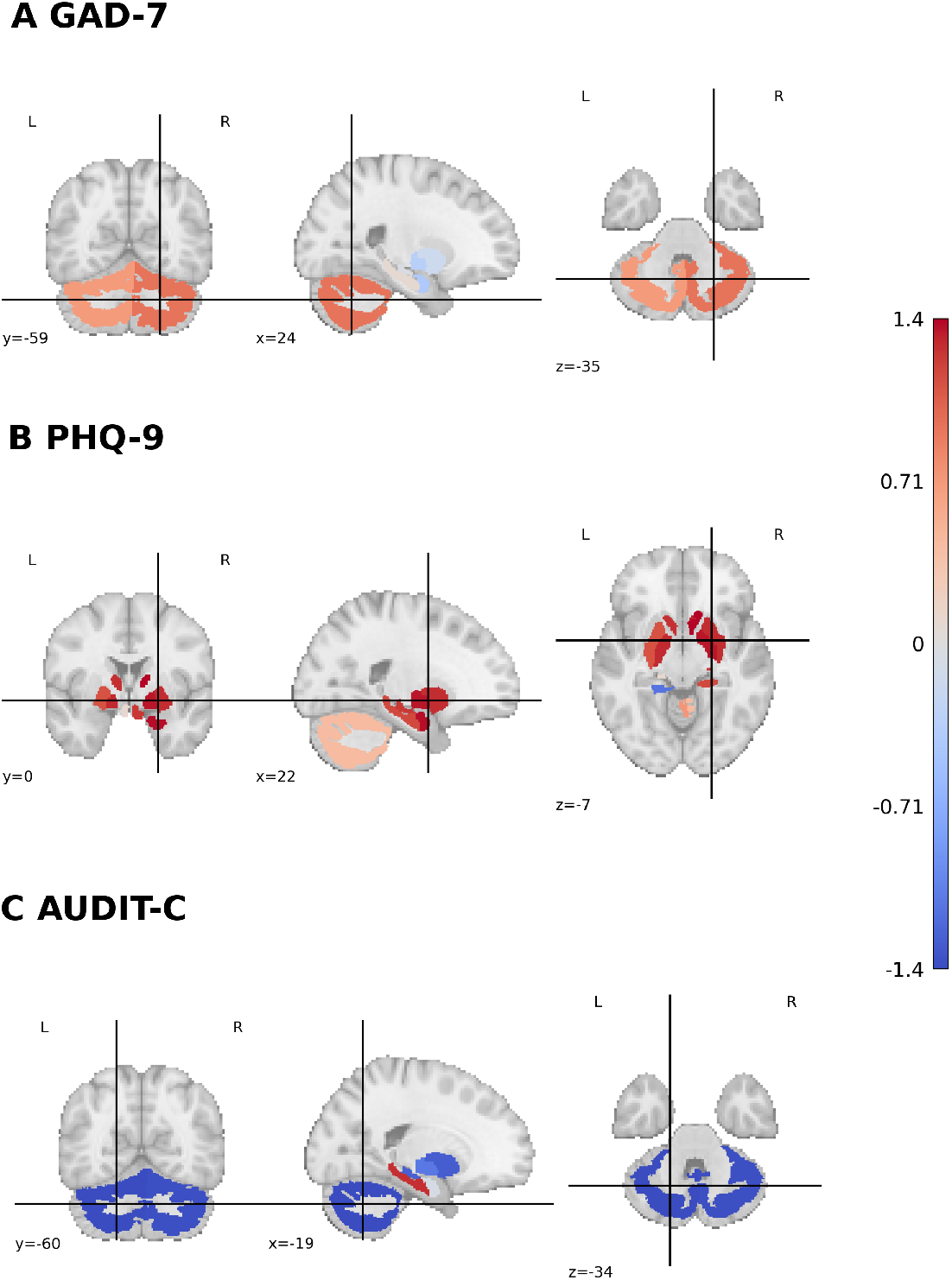
Z-scored associations between brain normative deviations and symptom scores for (A) GAD-7 (Generalized Anxiety Disorder-7), (B) PHQ-9 (Patient Health Questionnaire-9), and (C) AUDIT-C (Alcohol Use Disorders Identification Test–Consumption). Higher values reflect stronger regional associations with individual symptom expression. Associations are z-scored across all symptom scores and brain regions to emphasize relative patterns. These maps provide a heuristic summary of distributed associations between symptom-related deviation dimensions and regional GMV features, and should not be interpreted as maps of focal anatomical effects.

Direct correlations between deconfounded FreeSurfer GMVs and symptom scores yielded markedly different spatial patterns (Supplementary Figures 11 and 13), indicating that the normative deviation space captures diffuse structure not apparent in univariate region–symptom analyses.

### Classification Performance

Across symptom domains, models incorporating normative deviations consistently outperformed confounder-only baselines, with performance generally increasing with symptom severity (Table 2). Mean AUC and BACC generally increased with symptom severity, although variability also increased as group sizes decreased. Improvements were also observed for anxiety-related groups, although gains were smaller and less stable than for depression.

**Table 2:** Balanced accuracy (BACC) and area under the receiver operating characteristic curve (AUC) for all classifications. Values are mean ± standard deviation over 10-fold stratified crossvalidation, sorted by target. Targets are defined by symptom measures: GAD-7 (Generalized Anxiety Disorder-7), PHQ-9 (Patient Health Questionnaire-9), AUDIT-C (Alcohol Use Disorders Identification Test–Consumption), and ICD-10 diagnosis from hospital records or self-reported physician-reported diagnoses (diag.). Bold values indicate the best classifier for each target; in case of ties, the classifier with the smaller standard deviation is preferred, and if ties remain, the one requiring fewer features is chosen (Occam’s razor).

| Target | Confounders |  | + Deviations |  |
| --- | --- | --- | --- | --- |
|  | BACC (%) | AUC (%) | BACC (%) | AUC (%) |
| GAD-7 (2) | **58±2** | **61±2** | 58±2 | 61±2 |
| GAD-7 (3) | 58±6 | 62±5 | **61±2** | **64±3** |
| GAD-7 (4) | 59±9 | 62±12 | **65±8** | **65±6** |
| ANX ICD-10 | 55±3 | 60±4 | **62±2** | **63±4** |
| ANX (diag.) | 56±2 | 58±2 | **57±2** | **60±2** |
| PHQ-9 (1) | 58±1 | **61±1** | **59±2** | 61±1 |
| PHQ-9 (2) | 61±2 | 65±2 | **63±2** | **67±2** |
| PHQ-9 (3) | 59±6 | 65±7 | **61±6** | **69±7** |
| PHQ-9 (4) | 63±11 | 65±26 | **70±7** | **74±7** |
| MDD ICD-10 | 57±3 | 61±3 | **62±3** | **65±3** |
| MDD (diag.) | 58±1 | 61±1 | **59±1** | **62±1** |
| AUDIT-C (8) | **60±3** | **65±3** | 60±3 | 65±4 |
| AUDIT-C (9) | 63±3 | 68±4 | **64±4** | **69±3** |
| AUDIT-C $\geq 10$ | 63±3 | 68±3 | **66±3** | **72±3** |
| AUD ICD-10 | 55±9 | 56±10 | **62±4** | **63±4** |

In alcohol-related groups, models incorporating normative deviations again outperformed the confounder-only baseline, with performance increasing alongside symptom severity. It reached peak classification performance in the highest alcohol consumption group (AUC 72+/-3, BACC 66 +/-3). Using normative deviations yielded modest but consistent improvements over deconfounded FreeSurfer features across diagnostic groups (ΔBACC: median = 2%, mean = 2 *±* 2% SD; ΔAUC: median = 2%, mean = 1 *±* 2% SD; Supplementary Table 15). When restricting analyses to ICD-10–defined diagnostic groups, balanced accuracy increased further (ΔBACC *≈* +4–5%), with corresponding AUC gains for depressive and anxiety disorders (ΔAUC *≈* +3–4%), while alcohol consumption showed a small decrease in AUC (ΔAUC *≈ −* 1%). These results indicate that normative deviations carry information beyond demographic covariates, particularly at higher symptom severity.

## Discussion

In this study, we examined how depressive, anxiety-related, and alcohol-related symptoms relate to deviations in distributed gray matter structure in large population-based cohorts. We found that higher symptom severity was associated with greater deviation from normative structural variation. Depression and anxiety showed strongly overlapping deviation patterns, whereas alcohol-related symptoms deviated along a largely distinct direction, consistent with dimensional rather than diagnosis-specific organization.

Several choices in the normative modeling pipeline warrant consideration. We used MoCo-based contrastive representation learning to obtain compact features sensitive to distributed inter-individual structural differences, rather than optimizing voxelwise image reconstruction. This is particularly relevant for subtle psychiatric effects, where reconstruction-based objectives may prioritize dominant anatomical variance unrelated to symptoms. The 256-dimensional embedding size was chosen as a pragmatic representation size that balances representational capacity with downstream tractability and follows prior contrastive learning work using embeddings of this size [38, 39, 40]. The resulting normative model should therefore be interpreted as operating on a compact, data-driven structural representation rather than directly on voxelwise anatomy.

Several choices in the normative modeling pipeline warrant consideration. MoCo-based contrastive representation learning was used to obtain compact features sensitive to distributed interindividual structural differences rather than to optimize voxelwise image reconstruction. This is relevant for subtle psychiatric effects, where reconstruction objectives may prioritize dominant anatomical variance unrelated to symptoms. The 256-dimensional embedding size pragmatically balances representational capacity and downstream tractability, following prior contrastive learning work [38, 39, 40]. The normative model should therefore be interpreted as operating on a compact, data-driven structural representation rather than directly on voxelwise anatomy.

The directional overlap of MDD and GAD and specificity against AUD-related symptoms is consistent with the well-established clinical and biological overlap of MDD and GAD [46], including their high genetic correlation [47, 48] and the considerably lower overlap of either disorder with AUD. Importantly, this geometric organization should be interpreted in light of the contrastive representation learning stage used to derive the feature space. Contrastive objectives emphasize relative differences between individuals at the population level, rather than absolute regional abnormalities or deviations from an anatomical template. As a consequence, the resulting normative deviations reflect population-relative configurations of distributed brain structure rather than focal deficits. This representation bias may naturally favor dimensional and transdiagnostic organization, as variation is encoded along continuous axes of difference rather than partitioned into disorder-specific patterns. As some individuals with AUD show pronounced affective symptoms [49], we anticipated that such individuals would show an intermediate deviation profile. This was supported by our data, reinforcing the relevance of directional deviation analyses for capturing transdiagnostic neurobiological structure at the population level. The observed separation between internalizing and externalizing symptom profiles also aligns well with the dimensional structure proposed by the Hierarchical Taxonomy of Psychopathology (HiTOP) [50], which groups MDD and GAD within the internalizing spectrum and AUD within the externalizing domain, although it is important to note that some AUD-related alterations in brain structure may arise from the direct neurotoxic effects of alcohol rather than from an externalizing liability.

More broadly, and as a longer-term perspective, this approach may become useful for contextualizing psychiatric diagnoses as population cohorts increasingly provide imaging and phenotypic data across wider disorder spectra. Future analyses, beyond the scope of the present study, could extend this framework to additional psychiatric phenotypes, including psychotic and bipolar-spectrum conditions, and examine whether normative brain-structural deviation profiles correspond more closely to diagnostic categories, transdiagnostic spectra, or shared symptom dimensions. Analogous to genetic correlations revealing shared etiological architecture across disorders, including the close relationship between schizophrenia and bipolar disorder [51], population-scale normative imaging may help assess convergent or distinct deviation profiles and contribute to evaluating frameworks such as HiTOP, while avoiding the assumption that shared deviation geometry necessarily reflects shared etiology.

The observed improvements in individual-level prediction were concentrated at higher levels of symptom severity, particularly for depression. This pattern is consistent with the limited sensitivity of sMRI to subtle, heterogeneous alterations characteristic of common psychiatric conditions, and suggests that normative brain-structural deviations primarily enhance detection of more pronounced departures from the healthy population rather than enabling robust discrimination across the full symptom spectrum. When evaluated relative to conventional ROI-based GMV features, our normative deviations provided clearer benefits at the level of broader ICD-10–defined diagnostic groups. This likely reflects the ability of contrastive, normative representations to integrate distributed patterns of variation that are not well captured by region-wise measures. For ICD-10–defined AUD, divergence between BACC and AUC may be attributable to the smaller number of available cases (*N* = 192), highlighting the sensitivity of rank-based performance estimates to sample size in this context.

A post hoc mapping of deviation dimensions to regional GMV suggested that cerebellar regions contributed most consistently to symptom-associated deviation dimensions across domains. The strongest effects were observed for AUDIT-C and were negative in bilateral cerebellar cortex, indicating that higher alcohol-related symptom burden was linked to deviation dimensions associated with lower cerebellar GMV, consistent with prior evidence of cerebellar vulnerability in chronic alcohol use [52, 53]. Smaller, less pronounced contributions were also observed in subcortical regions, including the thalamus and caudate. In contrast, cortical contributions were weak and spatially distributed, with no dominant cortical region emerging as a focal contributor for depressive or anxiety-related symptoms. This lack of strong cortical localization is consistent with prior observations of distributed neural correlates in MDD [54] and psychiatric disorders in general [55], and suggests that affective-symptom-related deviations are better interpreted as multivariate, spatially distributed patterns than as isolated cortical abnormalities.

Several limitations should be considered. Temporal offsets between MRI acquisition and symptom assessment in UKB may attenuate brain–symptom associations, particularly for state-dependent symptoms; however, interval-stratified sensitivity analyses showed broadly stable group-level deviation directions when sufficient group sizes were retained. More generally, deviations were modest, heterogeneous, and primarily apparent at the group level, with limited individual-level classification performance. The analyses were restricted to sMRI and therefore do not capture functional, neurochemical, or molec- ular processes that may be more proximally related to symptoms. Moreover, representations were learned without direct symptom supervision, meaning that some symptom-relevant variation may not be encoded in the learned deviation space. Because Mahalanobis distances, shift estimates, and directional deviation vectors were defined relative to cohort-specific HC references, cross-cohort differences in absolute deviation magnitude or distance from the HC centroid are not directly interpretable. Region-level maps should also be interpreted cautiously, as the spatially unconstrained normative model was not optimized for anatomical localization and post hoc mappings provide indirect summaries; regional GMV features explained only a moderate fraction of variance in MoCo dimensions. Finally, although HCs were screened for low depressive, anxiety, and alcohol-related symptoms and available mood, anxiety, and alcohol-use diagnoses, the harmonized analysis did not allow comprehensive exclusion of other psychiatric conditions. This reflected the focus of available NAKO mental-health phenotypes as well as the unavailability of the required UKB diagnostic data for re-analysis during the revision period. Inclusion of individuals with unmeasured psychiatric conditions would likely increase normative heterogeneity and attenuate deviation estimates, particularly for MDD and GAD if these conditions share affective symptoms or structural correlates. As with all observational analyses, causal inferences cannot be drawn.

In summary, our analyses demonstrate that distributed normative deviations associated with depressive, anxiety, and alcohol-related symptoms show severity-dependent increases in magnitude and structured directional organization at the population level. Operating in a high-dimensional, image-derived representation space enabled the detection of subtle, spatially distributed deviation patterns. The overlap between depression- and anxiety-related deviations and their distinction from alcohol-related profiles is consistent with dimensional, transdiagnostic models of psychopathology. Although predictive gains are modest, the observed deviation axes may provide a way to stratify individuals according to continuous patterns of brain-structural variation that cut across conventional diagnostic categories. In large cohorts, such deviation-based representations may therefore be useful for identifying population-level neurobiological gradients of symptom burden, comorbidity, and severity rather than for making categorical individual-level diagnoses.

## Author Contributions

Julius Wiegert: Conceptualization, Methodology, Formal analysis, Data curation, Writing – original draft.

Sebastián Marty-Lombardi: Validation (code analysis), Writing – review & editing. Jailan Oweda: Data curation (imaging preprocessing).

Esra Lenz: Data curation (data extraction).

Antonia Mai, Amrou Abas, Xiuzhi Li: Data curation (imaging preprocessing support and genomic data processing).

Johannes Nitsche: Methodology (statistical analysis design), Writing – review & editing. Joonas Naamanka, Sebastian Volkmer, Andreas Meyer-Lindenberg: Writing – review & editing.

Tobias Gradinger, Fabian Streit, Urs Braun, Emanuel Schwarz: Conceptualization, Study design, Supervision.

Peter Ahnert, Klaus Berger, Hermann Brenner, Hans J. Grabe, Karin Halina Greiser, Johanna Klinger-König, André Karch, Michael Leitzmann, Claudia Meinke-Franz, Rafael Mikolajczyk, Thoralf Niendorf, Oliver Sander, Carsten Oliver Schmidt, Steffi G. Riedel-Heller, Annette Peters, Tobias Pischon: Resources (NAKO dataset), Writing – review & editing. Frauke Nees, Stephanie Witt, Kerstin Ritter, Josef Frank: Writing – review & editing.

All authors have reviewed and approved the manuscript.

## Acknowledgments

The project was conducted with data (NAKO-711) from the German National Cohort (NAKO) (http://www.nako.de/). The NAKO is funded by the Federal Ministry of Education and Research (BMBF) [project funding reference numbers: 01ER1301A/B/C, 01ER1511D, and 01ER1801A/B/C/D and 01ER2301A/B/C], federal states of Germany and the Helmholtz Association, the participating universities and the institutes of the Leibniz Association. We thank all participants who took part in the NAKO study and the staff of this research initiative. We also thank the participants and scientists involved in making the UK Biobank resource available (http://www.ukbiobank.ac.uk/). This study was conducted under UK Biobank application number 162313. The project was supported by the DZPG (German Centre for Mental Health Research) and by the BMBF (German Ministry of Education and Research) grant 01EE2303E. Fabian Streit is supported by a 2023 NARSAD Young Investigator Grant (#31537) from the Brain & Behavior Research Foundation with support from the Families for Borderline Personality Disorder Research. This work was supported by the Hector foundation II and was endorsed by German Center for Mental Health (DZPG).

## Data Availability

Access to and use of NAKO data and biosamples can be obtained via the electronic application portal (https://transfer.nako.de). Access to UK Biobank data requires application through the registration and application portal (http://ukbiobank.ac.uk/register-apply).

## Code Availability

All analysis were conducted in Python (version 3.12). All analysis scripts and the utilized packages are available via GitHub at https://github.com/wiegertj/DeepNormativeModeling.

