## Supplementary Material for "Breaking the Norm: Population-Scale Deviations of Brain Structure in Depression and Anxiety"

February 2026

*Corresponding author

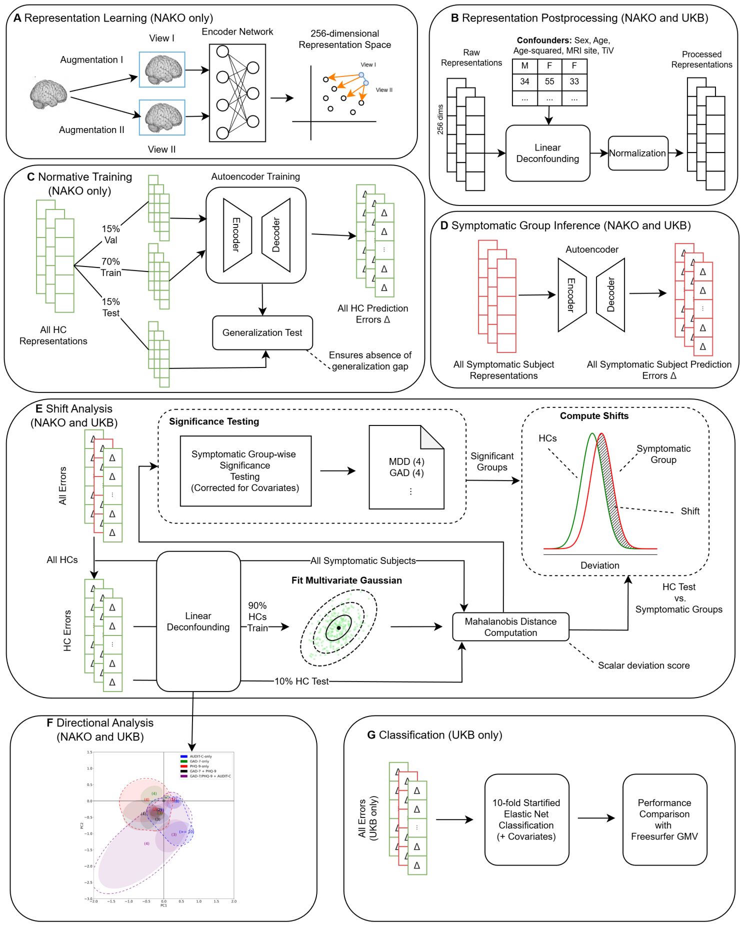


Figure 1: Complete study workflow. (**A**) Representation learning: brain MRI data were augmented and encoded into a 256-dimensional representation space using a contrastive learning framework. (**B**) Representation postprocessing: embeddings were linearly deconfounded for sex, age, age-squared, MRI site, and total intracranial volume (TIV), followed by normalization. (**C**) Normative training: an autoencoder was trained on healthy controls (HCs) with a train–validation–test split to predict HC representations, ensuring no generalization gap. (**D**) Symptomatic inference: deviations (Δ) were obtained for all symptomatic subjects by applying the HC-trained autoencoder. (**E**) Shift analysis: group-level deviations were compared against HC variability using multivariate Gaussian modeling, Mahalanobis distance, and bootstrap significance testing corrected for covariates. (**F**) Directional analysis: deviations were projected via principal component analysis (PCA). (**G**) Classification analysis in the UKB.

### 1 Data Filtering

#### 1.1 NAKO

| Field Name | Description | Missing Value Indicators | Affected |
|---|---|---|---|
| a_emo_miniscr_dp | MINI Screening MDD | 7777 | 226 |
| a_emo_phq9_kat | PHQ-9 category | 7777 | 1 240 |
| a_emo_gad7_dia | GAD-7 category | 7777 | 1 285 |
| a_alk_audit_c | AUDIT-C sum score | -99 | 1 285 |

Table 1: Missing values in key phenotypic fields from the NAKO cohort (German National Cohort). Field names correspond to raw database variables: `a_emo_miniscr_dp`, MINI International Neuropsychiatric Interview (MINI) screening for major depressive disorder (MDD); `a_emo_phq9_kat`, Patient Health Questionnaire-9 (PHQ-9) categorical score; `a_emo_gad7_dia`, Generalized Anxiety Disorder-7 (GAD-7) categorical score; `a_alk_audit_c`, Alcohol Use Disorders Identification Test–Consumption (AUDIT-C) sum score. “Missing value indicators” list the coding used for missing data in the raw dataset, and “Affected” gives the number of participants with missing entries.

#### 1.2 UKB

##### 1.2.1 Differing Standard Unit Definitions

In UK Biobank, alcohol units were defined as follows: pint or can of beer/lager/cider = 2 units; single shot of spirits (25 ml) = 1 unit; small glass of fortified wine = 1 unit; standard glass of wine (175 ml) = 2 units; large glass of wine (250 ml) = 3 units; bottle of wine (75 cl) = 9 units.

In NAKO, a standard drink corresponded to one small bottle or glass of beer (0.33 l), one small glass of wine or sparkling wine (0.125 l), or one simple shot of spirits (2 cl).

In general, the alcohol concentration per unit was lower in UKB compared to NAKO, which likely accounts for the higher proportion of participants with AUDIT-C $\geq$ 10 in UKB relative to NAKO (NAKO: 64% AUDIT 8, 25% AUDIT 9, 11% AUDIT $\geq$ 10; UKB: 40%, 29%, 30%).

##### 1.2.2 Neurodegenerative Exlcusion

| ICD-10 Code | Condition |
|---|---|
| G30 | Alzheimer's Disease |
| G20 | Parkinson's Disease |
| G10 | Huntington's Disease |
| G12.2 | Amyotrophic Lateral Sclerosis (ALS) |
| G35 | Multiple Sclerosis |
| I60–I64 | Stroke |
| G45.9 | Transient Ischemic Attack (TIA) |
| S06 | Traumatic Brain Injury (TBI) |
| M79.7 | Fibromyalgia |
| G25.0 | Essential Tremor |
| G24 | Dystonia |
| G61.0 | Guillain-Barré Syndrome |
| G36.0 | Neuromyelitis Optica |
| B20 | HIV-associated neurocognitive disorders |
| C71 | Brain tumor |
| G91 | Hydrocephalus |

Table 2: ICD-10 codes used to exclude participants with neurodegenerative, neurological, or related conditions from the UKB (UK Biobank) instance two MRI cohort. Conditions include major neurodegenerative diseases (e.g., Alzheimer's disease, Parkinson's disease, Huntington's disease, amyotrophic lateral sclerosis [ALS], multiple sclerosis), cerebrovascular events (stroke, transient ischemic attack [TIA]), traumatic brain injury (TBI), and other neurological disorders listed.

##### 1.2.3 Filtering Summary

| Step | Filtering Step | Remaining | Removed | Reason |
|---|---|---|---|---|
| 1 | Initial Sample | 43 248 | – | – |
| 2 | FreeSurfer QC | 39 369 | 3 879 | Any missing FreeSurfer statistics |
| 3 | GAD-7 Filter | 29 854 | 9 515 | Any missing GAD-7 question |
| 4 | PHQ-9 Filter | 25 977 | 3 877 | Any missing PHQ-9 question |
| 5 | AUDIT-C Filter | 25 977 | 0 | Any missing AUDIT-C question |

Table 3: Sequential filtering of the UKB (UK Biobank) MRI instance two cohort. "Remaining" indicates the number of participants retained after each step, and "Removed" gives the number excluded at that stage. FreeSurfer QC = exclusion of participants with missing FreeSurfer-derived brain structural measures; GAD-7 (Generalized Anxiety Disorder-7), PHQ-9 (Patient Health Questionnaire-9), and AUDIT-C (Alcohol Use Disorders Identification Test–Consumption) filters = exclusion of participants with missing responses to any item of the respective questionnaires.

##### 1.2.4 UKB Questionnaire Fields

| Field ID Showcase | Description | Missing Value Indicators |
|---|---|---|
| p120112 | PHQ-9 item | "Prefer not to answer" or *null* |
| p120111 | PHQ-9 item | "Prefer not to answer" or *null* |
| p120110 | PHQ-9 item | "Prefer not to answer" or *null* |
| p120109 | PHQ-9 item | "Prefer not to answer" or *null* |
| p120108 | PHQ-9 item | "Prefer not to answer" or *null* |
| p120107 | PHQ-9 item | "Prefer not to answer" or *null* |
| p120106 | PHQ-9 item | "Prefer not to answer" or *null* |
| p120105 | PHQ-9 item | "Prefer not to answer" or *null* |
| p120104 | PHQ-9 item | "Prefer not to answer" or *null* |

Table 4: UK Biobank field IDs corresponding to items of the PHQ-9 (Patient Health Questionnaire-9) depression scale. Each field represents a single questionnaire item. Missing values are coded as either "Prefer not to answer" or left as null entries in the dataset.

| **Field ID Showcase** | **Description** | **Missing Value Indicators** |
|---|---|---|
| p29058 | GAD-7 item | "Prefer not to answer" or *null* |
| p29059 | GAD-7 item | "Prefer not to answer" or *null* |
| p29060 | GAD-7 item | "Prefer not to answer" or *null* |
| p29061 | GAD-7 item | "Prefer not to answer" or *null* |
| p29062 | GAD-7 item | "Prefer not to answer" or *null* |
| p29063 | GAD-7 item | "Prefer not to answer" or *null* |
| p29064 | GAD-7 item | "Prefer not to answer" or *null* |

Table 5: UK Biobank field IDs corresponding to items of the GAD-7 (Generalized Anxiety Disorder-7) scale. Each field represents a single questionnaire item. Missing values are coded as either "Prefer not to answer" or left as null entries in the dataset.

| **Field ID** | **Description** | **Missing Value Indicators** |
|---|---|---|
| p20414 | Alcohol consumption frequency | "Prefer not to answer" or *null* |
| p20403 | Number of drinks consumed on a drinking day | "Prefer not to answer" or *null* |
| p29093 | Frequency of consuming six or more units of alcohol | "Prefer not to answer" or *null* |

Table 6: UK Biobank field IDs corresponding to items of the AUDIT-C (Alcohol Use Disorders Identification Test–Consumption). Each field represents one questionnaire item. Missing values are coded as either "Prefer not to answer" or left as null entries.

### 2 Technical Details

#### 2.1 Contrastive Feature Extraction

Normative modeling (NM) requires embedding brain magnetic resonance imaging (MRI) images into a suitable representational space that captures meaningful variations in brain structure. To achieve this, we use momentum contrast (MoCo) [1] for self-supervised feature extraction, reducing the dimensionality of grey matter volume (GMV) images to 256 dimensions.

MoCo implements contrastive learning by encouraging similar representations for augmented versions of the same image (positive pairs) while pushing apart representations of other images (negative pairs). It maintains a dynamic dictionary of encoded images (size 8 192) via a momentum encoder, ensuring a rich and diverse set of negative pairs, which is crucial for effective contrastive learning. By decoupling the number of negative pairs from batch size, MoCo remains scalable and efficient.

We evaluated three candidate embedding dimensionalities (128, 256, 512) during MoCo pretraining. Although a systematic optimization was not feasible due to the high computational cost of retraining MoCo with different settings—particularly because the augmentation pipeline is CPU-bound, cannot be precomputed, and requires on-the-fly random sampling—256 dimensions consistently yielded the smoothest and most stable loss convergence. In contrast, 128 and 512 dimensions showed slower or less stable convergence under otherwise identical settings. We further note that the optimal embedding size is not independent of other hyperparameters. In particular, the contrastive learning temperature and several architectural choices can interact with dimensionality to influence convergence and representation quality. Consequently, 256 dimensions was selected as a pragmatic choice balancing computational constraints and convergence stability. Furthermore, this dimensionality is also commonly used in prior contrastive learning work and balances representational capacity with downstream model complexity [2, 3, 4].

Unlike autoencoders (AEs), which prioritize input reconstruction, MoCo directly optimizes embedding similarity and dissimilarity, producing more structured representations. Additionally, it avoids the need for bottleneck tuning or reconstruction loss adjustments, making it a robust choice for feature extraction. Furthermore, in our experiments, AEs were unable to embed the data into meaningfully compact representations as effectively as MoCo. When constrained to smaller latent spaces, they either suffered from mode collapse or exhibited poor reconstruction quality.

To generate positive pairs, we apply two different random augmentations to the same original image, ensuring that both views retain the core anatomical features while introducing small variations. These augmentations—random affine transformations, random gamma adjustments, Gaussian noise, Gaussian blur, and random elastic deformations—help the model learn invariance to minor differences while still recognizing that both augmented views originate from the same underlying brain scan. We use the TorchIO library [5] to apply these transformations to the fully processed images.

Our MoCo encoder is a lightweight convolutional network implemented in PyTorch [6] consisting of five convolutional blocks, each followed by ReLU activation and max pooling (stride = 2) for downsampling, and three fully connected layers for gradual downsampling. The full architecture is detailed in Table 7.

| Block | Layer Type | Kernel Size | Channels | Stride | Activation |
|---|---|---|---|---|---|
| Block 1 | Conv3D | $3 \times 3 \times 3$ | $1 \rightarrow 32$ | 1 | ReLU |
| | MaxPool3D | $3 \times 3 \times 3$ | - | 2 | - |
| Block 2 | Conv3D | $3 \times 3 \times 3$ | $32 \rightarrow 32$ | 1 | ReLU |
| | MaxPool3D | $3 \times 3 \times 3$ | - | 2 | - |
| Block 3 | Conv3D | $3 \times 3 \times 3$ | $32 \rightarrow 64$ | 1 | ReLU |
| | MaxPool3D | $3 \times 3 \times 3$ | - | 2 | - |
| Block 4 | Conv3D | $3 \times 3 \times 3$ | $64 \rightarrow 64$ | 1 | ReLU |
| | MaxPool3D | $3 \times 3 \times 3$ | - | 2 | - |
| Block 5 | Conv3D | $3 \times 3 \times 3$ | $64 \rightarrow 64$ | 1 | ReLU |
| Fully Connected | Linear | - | $8000 \rightarrow 512$ | - | ReLU |
| | Linear | - | $512 \rightarrow 256$ | - | ReLU |
| | Linear | - | $256 \rightarrow 256$ | - | - |

Table 7: Architectural overview of the MoCo (Momentum Contrast) encoder convolutional neural network used for representation learning on 3D brain MRI data. Conv3D = three-dimensional convolutional layer; MaxPool3D = three-dimensional max-pooling layer; Linear = fully connected layer; ReLU = rectified linear unit activation function. Channels indicate input and output feature maps for each layer.

#### 2.2 Deconfounding Strategy

Many of the differences captured by MoCo appear to be influenced by confounding factors (Figure 2), which may obscure the underlying pathological signal. Therefore, we apply linear deconfounding using linear regression residualization with sex (one-hot encoded), age, age-squared, total intracranial volume (TiV), and recruitment center (one-hot encoded) as confounders. TiV was extracted using the TiV estimated by FreeSurfer [7]. We perform this linear deconfounding on each of the 256 MoCo dimensions. As shown in Figure 2, the magnitude of significant Spearman correlations between embeddings and confounders is markedly reduced after deconfounding, indicating effective removal of confounding effects.

We also explored more complex deconfounding strategies, such as conditional AEs and adversarial deconfounding, but found the optimization procedure to be diverging, likely due to the strong and pervasive nature of confounding in the dataset. As a result, we opted for rigorous linear deconfounding, potentially at the cost of some signal loss, with the aim of reducing—though not entirely eliminating—confounding bias in downstream analyses.

#### 2.3 Normative Modeling

We use the deconfounded MoCo embeddings as input for our NM pipeline, performing all modeling exclusively on the HC data. First, we min-max scale the embeddings to ensure stability during subsequent modeling. We then employ a simple 8-hidden-layer AE with ReLU activations to model normality of the deconfounded, scaled MoCo embeddings. Table 8 describes the AE architecture in detail.

The AE is trained to minimize the mean squared error (MSE) reconstruction loss. We use the AdamW optimizer (learning rate 0.0005) with batch size 64 for 300 epochs with early stopping based on validation loss (patience = 20), compressing the data into a latent representation of size

| Layer | Type | Output Dimension |
|---|---|---|
| Input | - | 256 |
| Block 1 | Linear | 150 |
| | ReLU | 150 |
| Block 2 | Linear | 100 |
| | ReLU | 100 |
| Block 3 | Linear | 75 |
| | ReLU | 75 |
| Block 4 | Linear | 50 |
| | ReLU | 50 |
| Block 5 | Linear | 75 |
| | ReLU | 75 |
| Block 6 | Linear | 100 |
| | ReLU | 100 |
| Block 7 | Linear | 150 |
| | ReLU | 150 |
| Block 8 | Linear | 256 |
| | Sigmoid | 256 |

Table 8: Architecture of the normative autoencoder used for reconstruction of deconfounded brain MRI embeddings. The network consists of eight fully connected (linear) blocks with symmetric compression and expansion, mapping the 256-dimensional input to a 50-dimensional bottleneck and back to 256 dimensions. ReLU = rectified linear unit activation function; Sigmoid = logistic activation function.

50, which was chosen due to its smallest generalization gap (see Table 9).

The generalization gap refers to the difference between a model's performance on training data versus unseen data. In NM, we want detected deviations to reflect meaningful pathological differences rather than artifacts of poor generalization. If a model has a large generalization gap, deviations in symptomatic subjects may be indistinguishable from errors due to poor generalization. By demonstrating that our model does not exhibit significantly larger errors on unseen HC (test set) than on seen HC (train and validation set), we increase the likelihood that deviations observed in symptomatic subjects truly reflect clinically meaningful effects. To confirm the absence of such effects, we compared reconstruction error distributions between training, validation, and test subsets of HCs using one-sided Mann–Whitney U tests. We found no significant differences between training and validation ($p = 0.26$) or between training and test ($p = 0.50$) reconstruction error distribution, supporting the stability and generalizability of our model.

#### 2.4 Shift Analysis

To quantify how diagnostic groups deviate from the natural variability present in HCs, we performed a shift analysis based on the full distribution of deviation scores. This approach moves beyond mean- or median-based comparisons by assessing how the *entire shape* of the deviation distribution in symptomatic groups diverges from that of HCs. What follows is a mathematical description of the steps performed to quantify the *shift*:

**1. Mahalanobis Distance Computation** For each subject, we computed the deviation from the normative reference distribution derived from HCs in the deconfounded embedding space. These distances reflect how strongly an individual's brain representation deviates from the normative manifold. After this step, we performed the group-wise significance testing described in the manuscript to ensure meaningfulness of the deviations.

**2. Kernel Density Estimation** For each diagnosis, we extracted the full set of deviations for all symptomatic groups and compared them to the HC distribution using Gaussian kernel density estimation (KDE) with Silverman's rule for bandwidth selection [8]. This procedure yields smooth estimates of the probability density functions for deviations in both HCs and symptomatic subjects.

**3. Exponential Weighting of Deviations** To prioritize clinically relevant patterns, we applied an exponential weighting function over the deviation axis, defined as:

$$w(x) = \exp\left(\frac{x}{\max(x)}\right),$$

where $x$ denotes the deviation. This weighting emphasizes larger deviations, under the assumption that they are more likely to reflect pathological alterations rather than normative variability, and we want the shift to be higher if it appears in high deviation density regions.

**4. Normalized Excess Area** We quantified the *weighted excess area*—the region where the symptomatic group density exceeds the HC density—according to:

$$\text{Excess Area} = \int_{x:\, f_{\text{symptomatic}}(x) > f_{\text{HC}}(x)} \left(f_{\text{symptomatic}}(x) - f_{\text{HC}}(x)\right) w(x)\, dx,$$

where $f_{\text{symptomatic}}(x)$ and $f_{\text{HC}}(x)$ are the KDEs of symptomatic and HC deviations, respectively, and $w(x)$ is the exponential weighting function.

The total weighted area under the symptomatic subject curve normalized this value:

$$\text{Normalization} = \int f_{\text{symptomatic}}(x) \cdot w(x)\, dx,$$

and the final metric was computed as:

$$\text{Fraction of Unexplained Abnormality (”shift”)} = \frac{\text{Excess Area}}{\text{Normalization}}.$$

This yields the final easy-to-interpret shift, emphasizing large deviations more harshly.

#### 2.5 Normative Model Stability

To evaluate the robustness of the estimated normative reference distribution parameters (mean $\mu$, covariance $\Sigma$), we conducted a bootstrap-based stability analysis.

Specifically, we resampled the full set of HC deviations with replacement 1,000 times, computing the mean vector and covariance matrix for each bootstrap sample. To assess variability, we used multiple stability metrics: (i) the average Euclidean distance and cosine similarity between bootstrap means and their overall centroid, and (ii) the average Frobenius norm between bootstrap covariance matrices and the mean covariance matrix. The Frobenius norm is defined as:

$$\|\Sigma\|_F = \sqrt{\sum_{i=1}^{d}\sum_{j=1}^{d}|a_{ij}|^2} \tag{1}$$

where $\Sigma \in \mathbb{R}^{d\times d}$ is the covariance matrix parameter of the Mahalanobis distance. This analysis was performed separately for both the NAKO and UKB cohorts.

In the NAKO cohort, results indicated low variability: the mean Euclidean distance between bootstrapped means was 0.0019, the cosine similarity was 0.9995, and the normalized covariance stability score was 0.0475 (with zero indicating perfect stability). The UKB cohort exhibited even greater stability, with a mean distance of 0.0002, a cosine similarity of 0.99998, and a normalized covariance score of 0.017. Covariance stability was defined as the average Frobenius norm between each bootstrap covariance and the mean covariance, normalized by the Frobenius norm of the mean covariance. This yields a scale-invariant measure of how consistently the covariance structure is preserved across resampled subsets, with lower values reflecting greater stability.

These findings confirm that the normative model Mahalanobis parameters (mean and covariance) are highly stable across bootstrap samples in both cohorts, supporting the reliability of the learned HC deviation distributions.

#### 2.6 Directional Analysis

**Algorithm 1**

**Require:**

Patient table with embeddings $X_p \in \mathbb{R}^{n_p \times D}$, confounders $C_p \in \mathbb{R}^{n_p \times q}$, and group indicators $\{g(i)\}_{i=1}^{n_p}$ over label set $\mathcal{G}$

HC table (already joined) with embeddings $X_h \in \mathbb{R}^{n_h \times D}$ and confounders $C_h \in \mathbb{R}^{n_h \times q}$

Concatenate $X \leftarrow \begin{bmatrix} X_p \\ X_h \end{bmatrix} \in \mathbb{R}^{N \times D}$, $C \leftarrow \begin{bmatrix} C_p \\ C_h \end{bmatrix} \in \mathbb{R}^{N \times q}$ where $N = n_p + n_h$.

**Linear residualization.** For each feature $j = 1, \dots, D$, solve

$$(\hat{\alpha}_j, \hat{\beta}_j) \in \arg\min_{\alpha,\beta} \left\| X_{\cdot j} - \alpha \mathbf{1} - C\beta \right\|_2^2, \quad R_{\cdot j} \leftarrow X_{\cdot j} - \hat{\alpha}_j \mathbf{1} - C\hat{\beta}_j.$$

Let $R \in \mathbb{R}^{N \times D}$ collect residuals.

**Standardize.** For $j = 1, \dots, D$ let $\mu_j = \frac{1}{N}\sum_{i=1}^{N} R_{ij}$ and $\sigma_j^2 = \frac{1}{N}\sum_{i=1}^{N}(R_{ij} - \mu_j)^2$. Set $\tilde{R}_{ij} = (R_{ij} - \mu_j)/\sigma_j$; denote $\tilde{R} \in \mathbb{R}^{N \times D}$.

**PCA to 2D.** Form $\Sigma = \frac{1}{N}\tilde{R}^\top \tilde{R}$ and take top-2 eigenvectors $W \in \mathbb{R}^{D \times 2}$. Project $Z = \tilde{R}W \in \mathbb{R}^{N \times 2}$ and split $Z = [Z_p; Z_h]$.

**HC robust center.** Compute $(c_h, \Lambda_h, Q_h) = \textsc{BootstrapEllipse}(Z_h, B)$ (Alg. 2).

**Recentering.** Translate $Z \leftarrow Z - \mathbf{1}c_h^\top$; obtain $Z_p, Z_h$ now centered at the HC median.

**Group ellipses.** For each label $g \in \mathcal{G}$, define $S_g = \{z_i \in Z_p : \ g(i) = g\}$. If $|S_g| > 0$, compute $(c_g, \Lambda_g, Q_g) = \textsc{BootstrapEllipse}(S_g, B)$.

**Ellipse parameters.** Each ellipse is represented by center $c \in \mathbb{R}^2$ and covariance-like shape $\Sigma^\star = Q\Lambda Q^\top$ where $\Lambda = \mathrm{diag}(\lambda_1, \lambda_2)$. The principal axes are $Qe_1, Qe_2$ with semi-axis lengths $a_k = \sqrt{\lambda_k}$.

**Algorithm 2** BootstrapEllipse: robust ellipse from geometric-median bootstraps

**Require:** Point set $S = \{x_i\}_{i=1}^{n} \subset \mathbb{R}^2$, bootstrap count $B$

**for** $b = 1$ to $B$ **do**

  Draw a bootstrap sample $S^{(b)}$ by sampling $n$ points with replacement from $S$

  Compute bootstrap geometric median

$$m_b \in \arg\min_{m \in \mathbb{R}^2} \sum_{x \in S^{(b)}} \|x - m\|_2 \quad \text{(e.g. via Weiszfeld's algorithm [9])}$$

**end for**

Form $M = \{m_b\}_{b=1}^{B}$ and its geometric median $c \in \arg\min_m \sum_{b=1}^{B} \|m_b - m\|_2$ (robust center).

Compute the covariance of $\{m_b\}$ with a small ridge: $\Sigma = \mathrm{Cov}(M) + \lambda I_2$ (e.g. $\lambda = 10^{-9}$).

Eigendecompose $\Sigma = Q\,\mathrm{diag}(\lambda_1, \lambda_2)\,Q^\top$ with $\lambda_1 \geq \lambda_2 \geq 0$.

**return** $c$ (center), $\Lambda = \mathrm{diag}(\lambda_1, \lambda_2)$, and $Q$ (principal directions).

#### 2.7 UKB External Validation Setup Details

In addition to transferring the models, we also transfer the deconfounding parameters for all confounders except the data collection center, as this variable reflects a dataset-specific effect. To address center-specific differences, we retrain the linear deconfounder for the center variable using only UKB data. For consistency in feature scaling, we apply the same min–max normalization used for the deconfounded embeddings. Due to cohort-specific effects, we had to re-estimate the Mahalanobis distance parameters, $\mu$ and $\Sigma$, from the HCs within the UKB cohort rather than transferring them from NAKO, because the original parameters yielded unrealistically high Mahalanobis distances in the UKB data (median 44.24 vs. 11.93 when using the NAKO parameters).

For estimating the Mahalanobis parameters used in the shift analysis and significance testing, we applied the same procedure as in NAKO. Specifically, we split the HC data into 90% training and 10% testing. The mean vector $\mu$ and covariance matrix $\Sigma$ were estimated on the 90% training set, and the shift analysis and significance testing were conducted on the remaining 10%. As detailed in Section 2.5, the Mahalanobis parameters in the UKB were found to be robust to sampling variability based on extensive bootstrapping, indicating that these parameters are highly stable within each cohort and largely insensitive to the specific subset of healthy controls used for fitting.

Finally, we reuse the PCA parameters fitted on the NAKO dataset for the directional deviation analysis in UKB.

### 3 Extended Results

#### 3.1 Autoencoder Dimensionality Tuning

Table 9: Evaluation of autoencoder bottleneck dimensionality. Models were trained with different bottleneck sizes and evaluated across five random seeds. Reported values are mean and standard deviation of validation reconstruction error (MSE) and downstream classification performance.

| Bottleneck | Val. MSE Mean | Val. SD |
|---:|---:|---:|
| 10 | 0.005560 | 0.000457 |
| 25 | 0.005228 | 0.001773 |
| 50 | 0.003839 | 0.000318 |
| 75 | 0.007397 | 0.003850 |
| 100 | 0.005952 | 0.003095 |

#### 3.2 Deconfounding

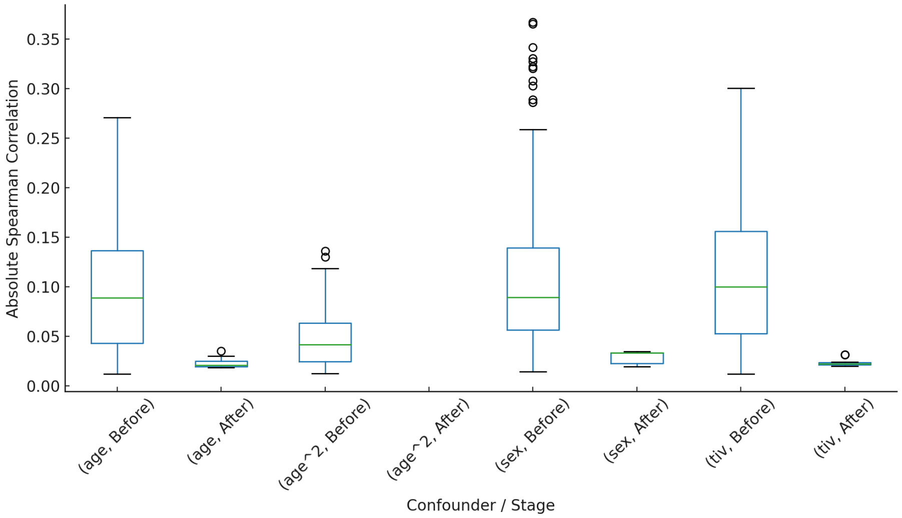


Figure 2: Distributions of absolute Spearman rank correlation coefficients between embedding dimensions and confounders before and after deconfounding. Confounders include age, age-squared, sex, and total intracranial volume (TIV). Only correlations significant at $p_{\mathrm{FDR}} < 0.05$ after Benjamini–Hochberg correction are shown.

#### 3.3 Shift Analysis Extended Results

Figure 3 presents the shift analysis using self-reported doctors' diagnoses and MINI-based diagnoses for NAKO, and ICD-10-based diagnoses for UKB.

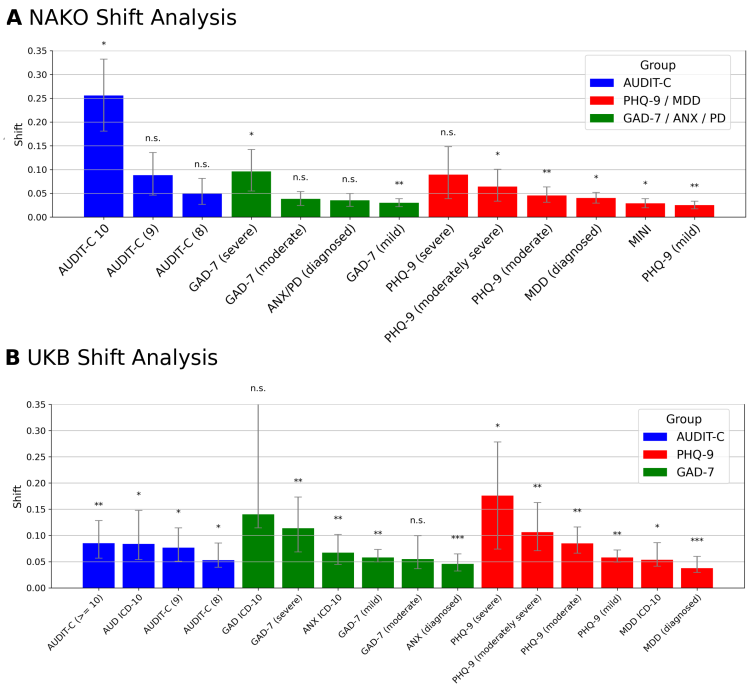


Figure 3: Fraction of group deviations unexplained by healthy control (HC) variability ("shift") for all symptomatic groups in the NAKO cohort (German National Cohort) (**A**) and the UKB cohort (UK Biobank) (**B**). Deviation scores were modeled using linear regression with group membership as the main predictor, adjusting for sex, age, age-squared, and sex–age interactions. Symptomatic groups were defined by AUDIT-C (Alcohol Use Disorders Identification Test–Consumption), PHQ-9 (Patient Health Questionnaire-9) for major depressive disorder (MDD), GAD-7 (Generalized Anxiety Disorder-7), and self-reported physician- or hospital record ICD-10–based diagnoses of anxiety disorders (ANX/PD) or MDD. Asterisks indicate significance levels after Benjamini–Hochberg false discovery rate (FDR) correction: * $p_{\mathrm{FDR}} < 0.05$, ** $p_{\mathrm{FDR}} < 0.01$, *** $p_{\mathrm{FDR}} < 0.005$.

Figure 4 presents the shift analysis in NAKO separated by sex.

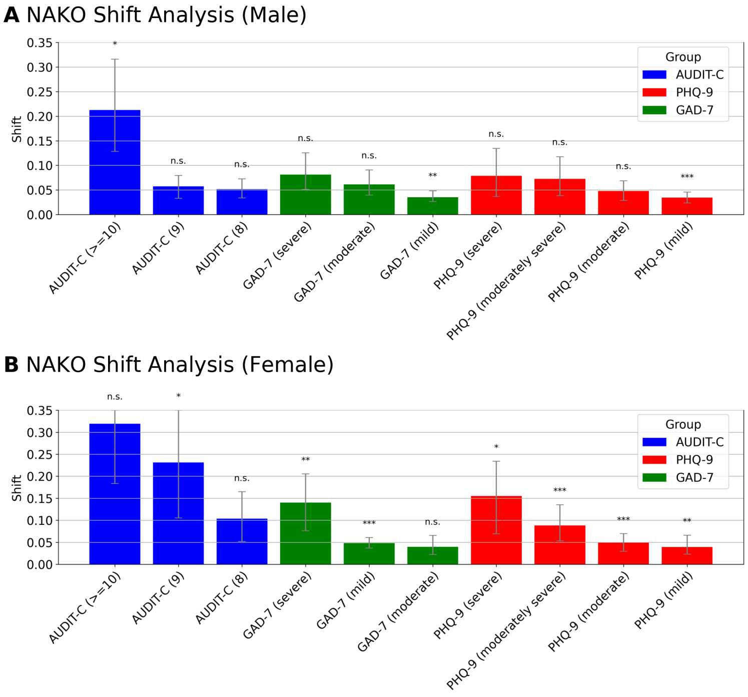


Figure 4: Fraction of group deviations unexplained by healthy control (HC) variability ("shift") in the NAKO cohort (German National Cohort), stratified by sex: (**A**) males and (**B**) females. Deviation scores were modeled using linear regression with group membership as the main predictor, adjusting for age and age-squared. Symptomatic groups were defined by AUDIT-C (Alcohol Use Disorders Identification Test–Consumption), PHQ-9 (Patient Health Questionnaire-9), and GAD-7 (Generalized Anxiety Disorder-7) thresholds. Asterisks indicate significance levels after Benjamini–Hochberg false discovery rate (FDR) correction: * $p_{\mathrm{FDR}} < 0.05$, ** $p_{\mathrm{FDR}} < 0.01$, *** $p_{\mathrm{FDR}} < 0.005$.

| **Diagnosis** | **p-value** | $\mathbf{p}_{\mathrm{FDR}}$ |
|---|---|---|
| MINI | 0.0080 | 0.0174 |
| MDD (diagnosed) | 0.0070 | 0.0174 |
| ANX/PD (diagnosed) | 0.2517 | 0.2975 |
| AUDIT-C (8) | 0.7128 | 0.7128 |
| AUDIT-C (9) | 0.0857 | 0.1237 |
| AUDIT-C (>=10) | 0.0045 | 0.0147 |
| PHQ-9 (mild) | 0.0016 | 0.0088 |
| PHQ-9 (moderate) | 0.0008 | 0.0088 |
| PHQ-9 (moderately severe) | 0.0143 | 0.0265 |
| PHQ-9 (severe) | 0.1537 | 0.1998 |
| GAD-7 (mild) | 0.0020 | 0.0088 |
| GAD-7 (moderate) | 0.4490 | 0.4864 |
| GAD-7 (severe) | 0.0258 | 0.0419 |

Table 10: ANOVA results for the NAKO cohort (German National Cohort), reporting raw p-values (test of the null hypothesis that group means are equal, PR($> F$)) and Benjamini–Hochberg false discovery rate (FDR)–adjusted p-values ($p_{\mathrm{FDR}}$) for each diagnostic group. MINI = Mini International Neuropsychiatric Interview; MDD = major depressive disorder; ANX/PD = anxiety or panic disorder (doctor's diagnosis); AUDIT-C = Alcohol Use Disorders Identification Test–Consumption; PHQ-9 = Patient Health Questionnaire-9; GAD-7 = Generalized Anxiety Disorder-7.

| **Tested Group** | **p-value** | $\mathbf{p}_{\text{FDR}}$ |
|---|---|---|
| MDD (ICD-10) | 0.0153 | 0.0231 |
| AUD (ICD-10) | 0.0396 | 0.0453 |
| AUDIT-C (8) | 0.0391 | 0.0453 |
| AUDIT-C (9) | 0.0173 | 0.0231 |
| AUDIT-C (>=10) | 0.0054 | 0.0096 |
| ANX (ICD-10) | 0.0025 | 0.0071 |
| GAD (ICD-10) | 0.2325 | 0.2874 |
| PHQ-9 (mild) | 0.0031 | 0.0071 |
| PHQ-9 (moderate) | 0.0010 | 0.0052 |
| PHQ-9 (moderately severe) | 0.0025 | 0.0071 |
| PHQ-9 (severe) | 0.0166 | 0.0231 |
| GAD-7 (mild) | 0.0031 | 0.0071 |
| GAD-7 (moderate) | 0.1723 | 0.1838 |
| GAD-7 (severe) | 0.0052 | 0.0096 |
| MDD (diagnosed) | 0.0002 | 0.0015 |
| ANX (diagnosed) | 0.00005 | 0.0008 |

Table 11: ANOVA results for the UKB cohort (UK Biobank), reporting raw p-values (test of the null hypothesis that group means are equal, PR($> F$)) and Benjamini–Hochberg false discovery rate (FDR)–adjusted p-values ($p_{\text{FDR}}$) for each tested group. MDD = major depressive disorder; AUD = alcohol use disorder; ANX = anxiety disorder; GAD = generalized anxiety disorder; PHQ-9 = Patient Health Questionnaire-9; GAD-7 = Generalized Anxiety Disorder-7.

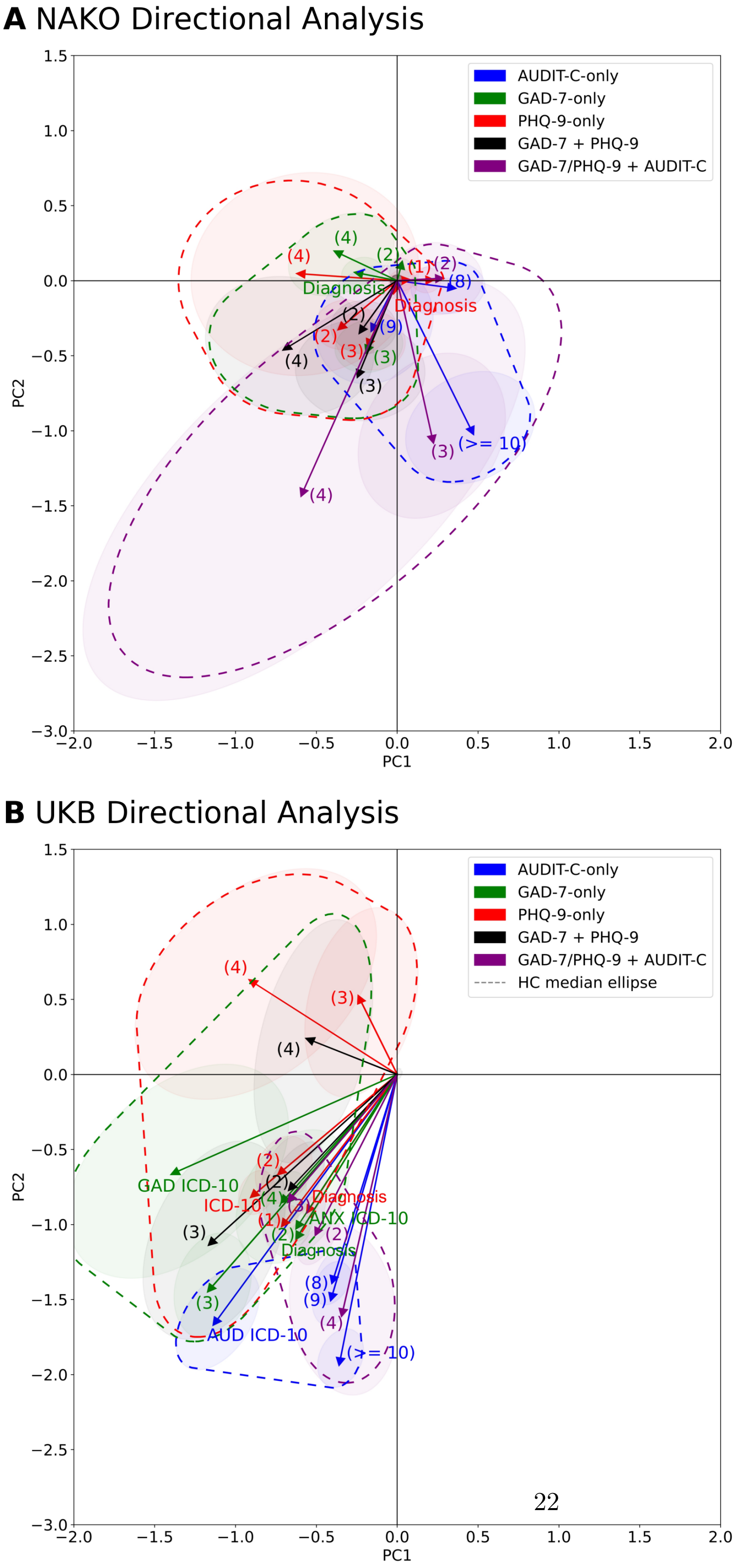


Figure 5: Directional analyses of normative deviation vectors based on principal component analysis (PCA; two dimensions, 21% explained variance) for diagnostic groups in the NAKO (German National Cohort) (**A**) and UKB (UK Biobank) cohort (**B**). Shaded ellipses show the ±1 SD contour of the bootstrap (1,000 resamples) distribution around the group-level geometric median vectors. The dark ellipse at the origin represents the median deviation among healthy controls (HCs), serving as a reference for normative variability. Numbers indicate severity levels: PHQ-9 (Patient Health Questionnaire-9) levels (1–4) = mild, moderate, moderately severe, severe; GAD-7 (Generalized Anxiety Disorder-7) levels (2–4) = mild, moderate, severe; AUDIT-C (Alcohol Use Disorders Identification Test–Consumption) = symptom count. "MDD/GAD + ALC" denotes individuals with acute major depressive disorder (MDD) or generalized anxiety disorder (GAD) symptoms and elevated alcohol use (AUDIT-C $\geq$ 10). "ICD-10" refers to International Classification of Diseases, 10th Revision, hospital record codes, and "Diagnosis" to self-reported conditions. Red ellipses = PHQ/GAD groups, blue = AUDIT-C groups, purple = comorbid groups.

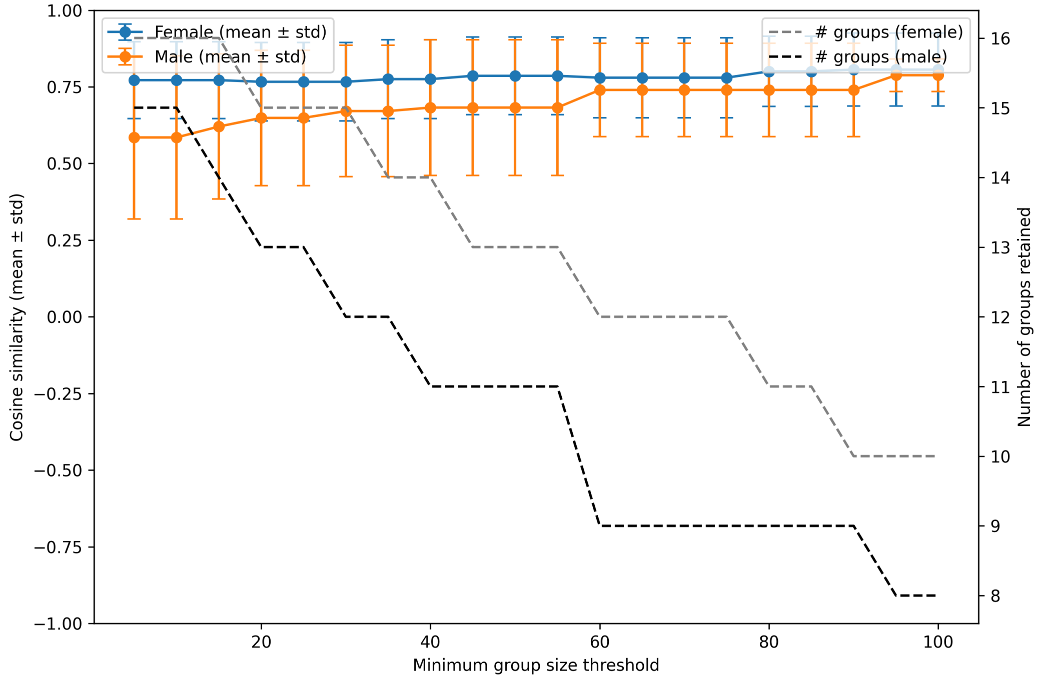


Figure 6: Cosine similarity analysis results for directional analysis with sex stratification. Error bars denote standard deviations across groups; similarity is computed to the joint-sex median vector. The x-axis indicates the minimum group size required for inclusion in the analysis. Dashed lines show the number of groups meeting this threshold.

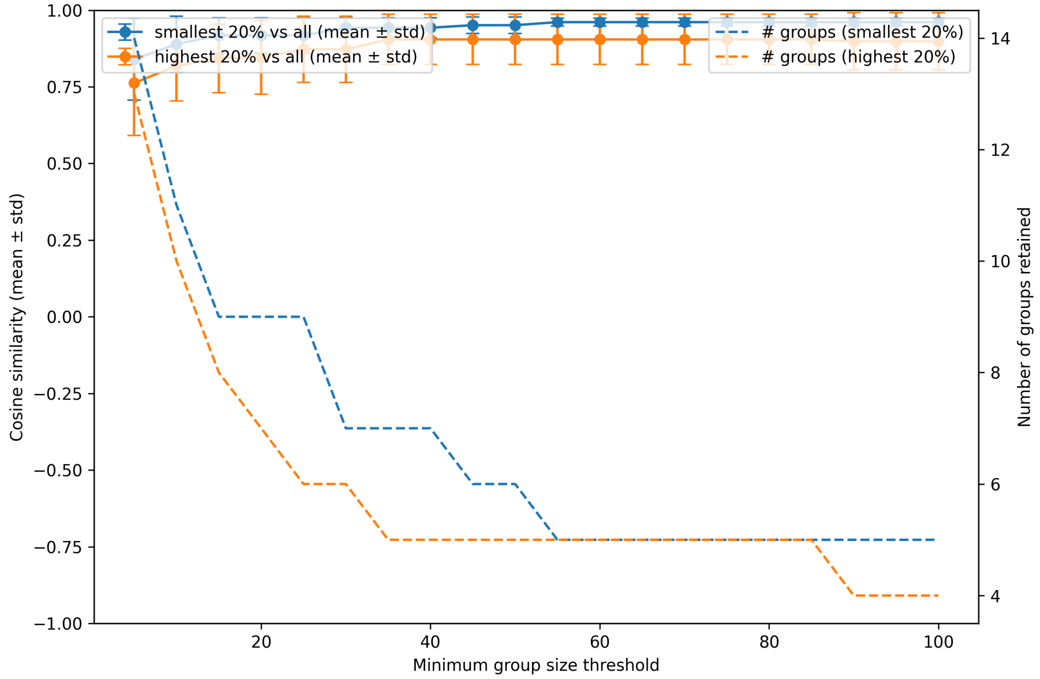


Figure 7: Influence of the interval between MRI and questionnaire acquisition. Error bars denote the standard deviation across groups. Cosine similarity (y-axis) reflects the similarity of each group's median vector (bottom 20% and top 20% of the interval distribution) to the unfiltered median vector. The x-axis indicates the minimum group size required for inclusion in the analysis. Dashed lines show the number of groups meeting this threshold.

##### 3.4 List of FreeSurfer Measures

Table 12: List of Freesurfer measures used for deviation association analysis and classification baseline

| **FreeSurfer Measure** |
|---|
| aseg.volume_Left-Cerebellum-Cortex |
| aseg.volume_Left-Thalamus |
| aseg.volume_Left-Caudate |
| aseg.volume_Left-Putamen |
| aseg.volume_Left-Pallidum |
| aseg.volume_Left-Hippocampus |
| aseg.volume_Left-Amygdala |
| aseg.volume_Left-Accumbens-area |
| aseg.volume_Left-VentralDC |
| aseg.volume_Right-Cerebellum-Cortex |
| aseg.volume_Right-Thalamus |
| aseg.volume_Right-Caudate |
| aseg.volume_Right-Putamen |
| aseg.volume_Right-Pallidum |
| aseg.volume_Right-Hippocampus |
| aseg.volume_Right-Amygdala |
| aseg.volume_Right-Accumbens-area |
| aseg.volume_Right-VentralDC |
| aseg.volume_non-WM-hypointensities |
| aseg.volume_Left-non-WM-hypointensities |
| aseg.volume_Right-non-WM-hypointensities |
| aseg.volume_BrainSegVol |
| aseg.volume_BrainSegVolNotVent |
| aseg.volume_lhCortexVol |
| aseg.volume_rhCortexVol |
| aseg.volume_CortexVol |
| aseg.volume_SubCortGrayVol |
| aseg.volume_TotalGrayVol |
| aseg.volume_SupraTentorialVol |
| aseg.volume_SupraTentorialVolNotVent |
| aseg.volume_EstimatedTotalIntraCranialVol |
| lh.aparc.volume_lh_bankssts_volume |
| lh.aparc.volume_lh_caudalanteriorcingulate_volume |
| lh.aparc.volume_lh_caudalmiddlefrontal_volume |
| lh.aparc.volume_lh_cuneus_volume |
| lh.aparc.volume_lh_entorhinal_volume |
| lh.aparc.volume_lh_fusiform_volume |
| lh.aparc.volume_lh_inferiorparietal_volume |
| lh.aparc.volume_lh_inferiortemporal_volume |
| lh.aparc.volume_lh_isthmuscingulate_volume |

| **FreeSurfer Measure (continued)** |
|---|
| lh.aparc.volume_lh_lateraloccipital_volume |
| lh.aparc.volume_lh_lateralorbitofrontal_volume |
| lh.aparc.volume_lh_lingual_volume |
| lh.aparc.volume_lh_medialorbitofrontal_volume |
| lh.aparc.volume_lh_middletemporal_volume |
| lh.aparc.volume_lh_parahippocampal_volume |
| lh.aparc.volume_lh_paracentral_volume |
| lh.aparc.volume_lh_parsopercularis_volume |
| lh.aparc.volume_lh_parsorbitalis_volume |
| lh.aparc.volume_lh_parstriangularis_volume |
| lh.aparc.volume_lh_pericalcarine_volume |
| lh.aparc.volume_lh_postcentral_volume |
| lh.aparc.volume_lh_posteriorcingulate_volume |
| lh.aparc.volume_lh_precentral_volume |
| lh.aparc.volume_lh_precuneus_volume |
| lh.aparc.volume_lh_rostralanteriorcingulate_volume |
| lh.aparc.volume_lh_rostralmiddlefrontal_volume |
| lh.aparc.volume_lh_superiorfrontal_volume |
| lh.aparc.volume_lh_superiorparietal_volume |
| lh.aparc.volume_lh_superiortemporal_volume |
| lh.aparc.volume_lh_supramarginal_volume |
| lh.aparc.volume_lh_frontalpole_volume |
| lh.aparc.volume_lh_temporalpole_volume |
| lh.aparc.volume_lh_transversetemporal_volume |
| lh.aparc.volume_lh_insula_volume |
| rh.aparc.volume_rh_bankssts_volume |
| rh.aparc.volume_rh_caudalanteriorcingulate_volume |
| rh.aparc.volume_rh_caudalmiddlefrontal_volume |
| rh.aparc.volume_rh_cuneus_volume |
| rh.aparc.volume_rh_entorhinal_volume |
| rh.aparc.volume_rh_fusiform_volume |
| rh.aparc.volume_rh_inferiorparietal_volume |
| rh.aparc.volume_rh_inferiortemporal_volume |
| rh.aparc.volume_rh_isthmuscingulate_volume |
| rh.aparc.volume_rh_lateraloccipital_volume |
| rh.aparc.volume_rh_lateralorbitofrontal_volume |
| rh.aparc.volume_rh_lingual_volume |
| rh.aparc.volume_rh_medialorbitofrontal_volume |
| rh.aparc.volume_rh_middletemporal_volume |
| rh.aparc.volume_rh_parahippocampal_volume |
| rh.aparc.volume_rh_paracentral_volume |
| rh.aparc.volume_rh_parsopercularis_volume |
| rh.aparc.volume_rh_parsorbitalis_volume |
| rh.aparc.volume_rh_parstriangularis_volume |
| rh.aparc.volume_rh_pericalcarine_volume |

| **FreeSurfer Measure (continued)** |
|---|
| rh.aparc.volume_rh_postcentral_volume |
| rh.aparc.volume_rh_posteriorcingulate_volume |
| rh.aparc.volume_rh_precentral_volume |
| rh.aparc.volume_rh_precuneus_volume |
| rh.aparc.volume_rh_rostralanteriorcingulate_volume |
| rh.aparc.volume_rh_rostralmiddlefrontal_volume |
| rh.aparc.volume_rh_superiorfrontal_volume |
| rh.aparc.volume_rh_superiorparietal_volume |
| rh.aparc.volume_rh_superiortemporal_volume |
| rh.aparc.volume_rh_supramarginal_volume |
| rh.aparc.volume_rh_frontalpole_volume |
| rh.aparc.volume_rh_temporalpole_volume |
| rh.aparc.volume_rh_transversetemporal_volume |
| rh.aparc.volume_rh_insula_volume |

#### 3.5 Correlation Analysis

Table 13: Significant Spearman rank correlations between latent embedding dimensions and symptom severity scores after false discovery rate (FDR) correction ($q < 0.05$). The estimated proportion of true null hypotheses ($\pi_0$), derived using the q-value procedure [10], was 0.74. For each association, Spearman's $r$, uncorrected $p$-values, and corresponding FDR-adjusted $q$-values are reported.

| Score | Dimension | Spearman r | $q_{\text{value}}$ | $p_{\text{value\_uncorrected}}$ |
|---|---|---|---|---|
| AUDIT-C sum score | 48 | -0.027 | 1.135e-03 | 3.738e-06 |
| AUDIT-C sum score | 68 | -0.027 | 1.135e-03 | 4.027e-06 |
| AUDIT-C sum score | 221 | 0.026 | 1.982e-03 | 1.055e-05 |
| AUDIT-C sum score | 20 | -0.025 | 2.174e-03 | 1.691e-05 |
| AUDIT-C sum score | 136 | 0.025 | 2.174e-03 | 1.929e-05 |
| AUDIT-C sum score | 13 | -0.024 | 3.245e-03 | 3.582e-05 |
| AUDIT-C sum score | 142 | 0.024 | 3.245e-03 | 4.605e-05 |
| AUDIT-C sum score | 145 | -0.024 | 3.245e-03 | 4.563e-05 |
| AUDIT-C sum score | 114 | 0.024 | 3.498e-03 | 5.585e-05 |
| AUDIT-C sum score | 213 | 0.024 | 3.793e-03 | 6.729e-05 |
| AUDIT-C sum score | 16 | -0.023 | 3.817e-03 | 7.449e-05 |
| AUDIT-C sum score | 217 | 0.023 | 5.072e-03 | 1.260e-04 |
| AUDIT-C sum score | 207 | 0.022 | 7.509e-03 | 2.132e-04 |
| AUDIT-C sum score | 150 | 0.021 | 9.336e-03 | 2.982e-04 |
| AUDIT-C sum score | 227 | -0.021 | 9.336e-03 | 2.837e-04 |
| AUDIT-C sum score | 52 | 0.021 | 9.996e-03 | 3.370e-04 |
| AUDIT-C sum score | 100 | 0.021 | 1.120e-02 | 4.174e-04 |

Continued on next page

Table 13: Significant Spearman rank correlations between latent embedding dimensions and symptom severity scores after false discovery rate (FDR) correction ($q < 0.05$). The estimated proportion of true null hypotheses ($\pi_0$), derived using the q-value procedure [10], was 0.74. For each association, Spearman's $r$, uncorrected $p$-values, and corresponding FDR-adjusted $q$-values are reported.

| Score | Dimension | Spearman r | $q_{\text{value}}$ | $p_{\text{value\_uncorrected}}$ |
|---|---|---|---|---|
| AUDIT-C sum score | 224 | -0.021 | 1.120e-02 | 4.165e-04 |
| AUDIT-C sum score | 162 | 0.021 | 1.135e-02 | 4.553e-04 |
| AUDIT-C sum score | 204 | -0.021 | 1.135e-02 | 4.631e-04 |
| AUDIT-C sum score | 155 | 0.020 | 1.155e-02 | 5.326e-04 |
| AUDIT-C sum score | 86 | -0.020 | 1.268e-02 | 6.074e-04 |
| AUDIT-C sum score | 73 | 0.020 | 1.386e-02 | 6.884e-04 |
| AUDIT-C sum score | 62 | 0.019 | 2.478e-02 | 1.599e-03 |
| AUDIT-C sum score | 87 | 0.019 | 2.478e-02 | 1.528e-03 |
| AUDIT-C sum score | 122 | 0.019 | 2.478e-02 | 1.581e-03 |
| AUDIT-C sum score | 216 | -0.019 | 2.478e-02 | 1.533e-03 |
| AUDIT-C sum score | 182 | 0.018 | 2.900e-02 | 1.955e-03 |
| AUDIT-C sum score | 102 | 0.018 | 3.010e-02 | 2.083e-03 |
| AUDIT-C sum score | 112 | 0.018 | 3.104e-02 | 2.238e-03 |
| AUDIT-C sum score | 223 | -0.017 | 3.931e-02 | 3.208e-03 |
| AUDIT-C sum score | 31 | -0.017 | 4.784e-02 | 4.244e-03 |
| AUDIT-C sum score | 177 | -0.017 | 4.784e-02 | 4.161e-03 |
| AUDIT-C sum score | 61 | 0.017 | 4.888e-02 | 4.509e-03 |
| AUDIT-C sum score | 173 | 0.017 | 4.901e-02 | 4.609e-03 |
| GAD-7 sum score | 100 | -0.019 | 2.478e-02 | 1.306e-03 |
| GAD-7 sum score | 241 | 0.019 | 2.478e-02 | 1.445e-03 |
| GAD-7 sum score | 217 | -0.017 | 3.931e-02 | 3.087e-03 |
| GAD-7 sum score | 58 | 0.017 | 4.821e-02 | 4.362e-03 |
| PHQ-9 sum score | 76 | 0.023 | 4.865e-03 | 1.069e-04 |
| PHQ-9 sum score | 217 | -0.023 | 4.865e-03 | 1.122e-04 |
| PHQ-9 sum score | 246 | 0.022 | 6.592e-03 | 1.754e-04 |
| PHQ-9 sum score | 100 | -0.021 | 1.155e-02 | 4.924e-04 |
| PHQ-9 sum score | 205 | 0.020 | 1.155e-02 | 5.170e-04 |
| PHQ-9 sum score | 132 | -0.019 | 2.478e-02 | 1.626e-03 |
| PHQ-9 sum score | 225 | 0.019 | 2.478e-02 | 1.418e-03 |
| PHQ-9 sum score | 231 | 0.019 | 2.478e-02 | 1.606e-03 |
| PHQ-9 sum score | 6 | -0.018 | 3.104e-02 | 2.313e-03 |
| PHQ-9 sum score | 233 | 0.018 | 3.104e-02 | 2.267e-03 |
| PHQ-9 sum score | 45 | -0.017 | 3.931e-02 | 3.203e-03 |
| PHQ-9 sum score | 91 | -0.017 | 3.931e-02 | 3.126e-03 |
| PHQ-9 sum score | 227 | 0.017 | 4.161e-02 | 3.470e-03 |
| PHQ-9 sum score | 216 | -0.017 | 4.784e-02 | 4.222e-03 |

##### 3.6 z-Scored Symptom Score Mapping Brains

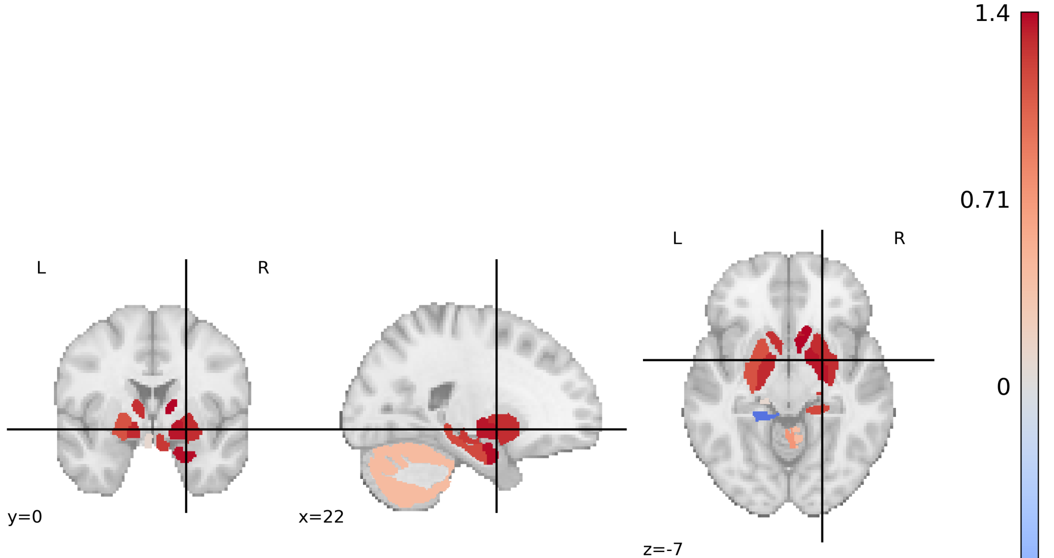


Figure 8: Z-scored heuristic associations between latent embedding dimensions and PHQ-9 (Patient Health Questionnaire-9) depression sum scores, mapped onto the MNI-152 (Montreal Neurological Institute) brain template. Higher z-scores indicate stronger relative associations across brain regions.

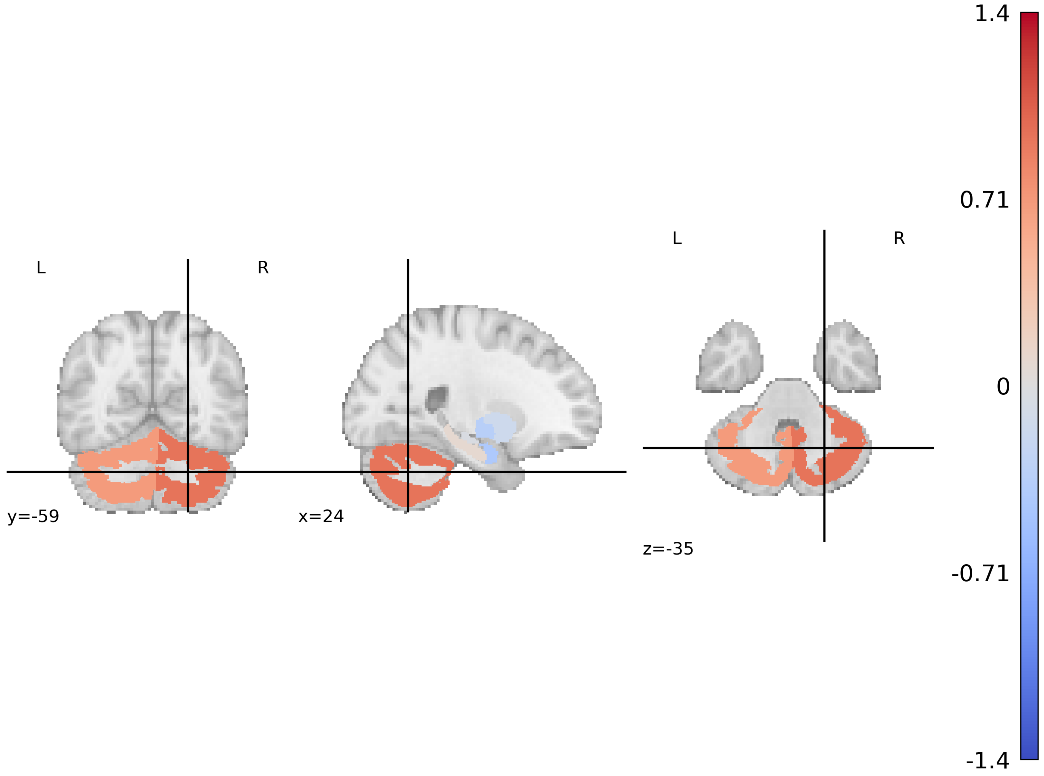


Figure 9: Z-scored heuristic associations between latent embedding dimensions and GAD-7 (Generalized Anxiety Disorder-7) anxiety sum scores, mapped onto the MNI-152 (Montreal Neurological Institute) brain template. Higher z-scores indicate stronger relative associations across brain regions.

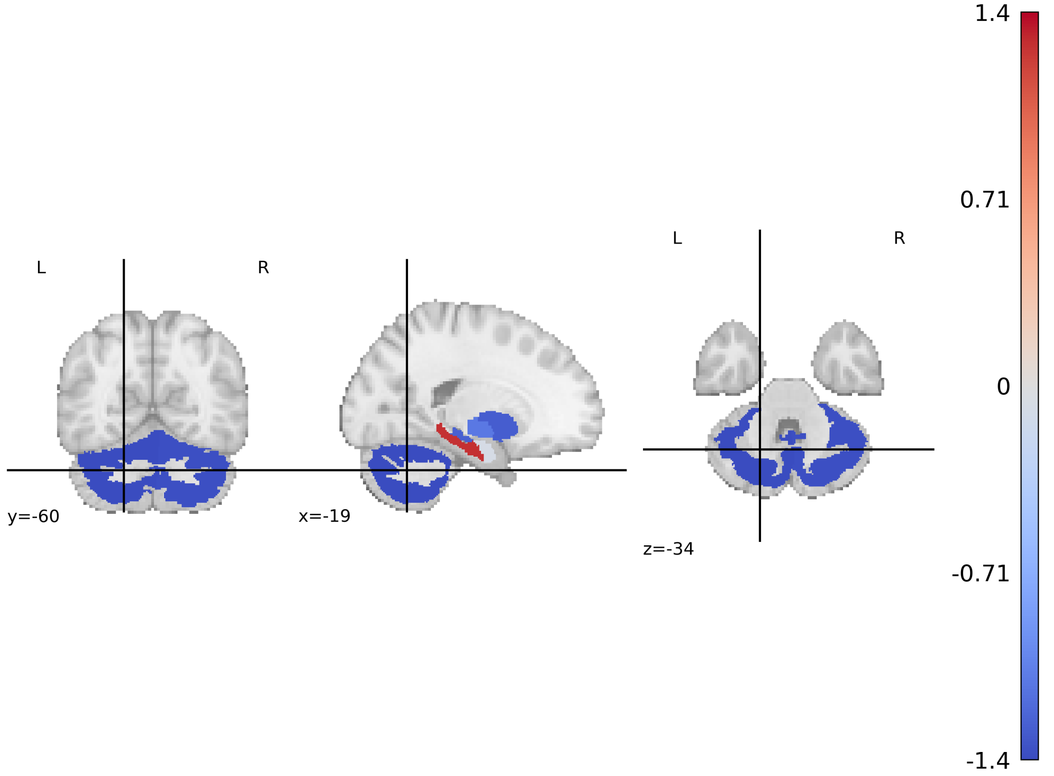


Figure 10: Z-scored heuristic associations between latent embedding dimensions and AUDIT-C (Alcohol Use Disorders Identification Test–Consumption) alcohol use sum scores, mapped onto the MNI-152 (Montreal Neurological Institute) brain template. Higher z-scores indicate stronger relative associations across brain regions.

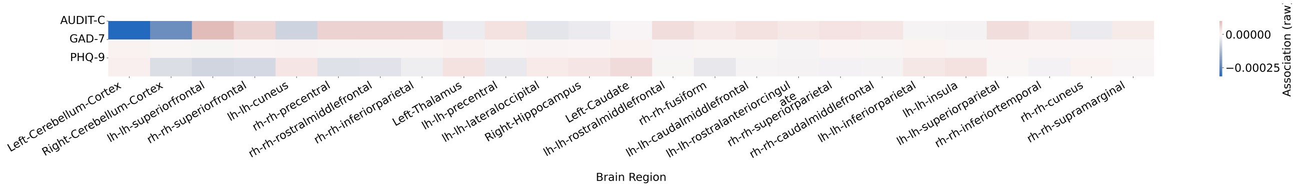


Figure 11: Top 25 brain regions ranked by absolute effect size of heuristic associations between latent embedding dimensions and symptom severity scores. Rows correspond to AUDIT-C (Alcohol Use Disorders Identification Test–Consumption), GAD-7 (Generalized Anxiety Disorder-7), and PHQ-9 (Patient Health Questionnaire-9). Columns indicate FreeSurfer-derived brain regions. Colors denote the relative strength and direction of association (blue = negative, red = positive), with the scale representing association strength.

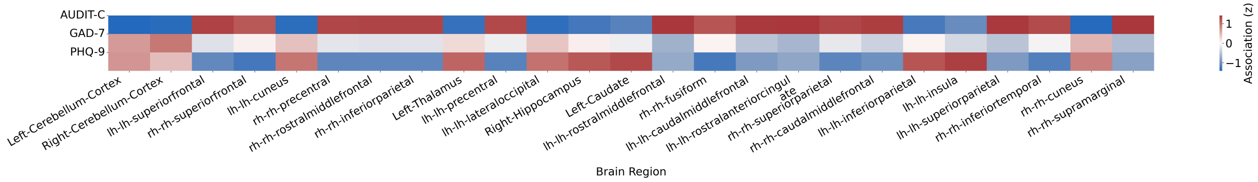


Figure 12: Top 25 brain regions ranked by absolute effect size of z-scored heuristic associations between latent embedding dimensions and symptom severity scores. Rows correspond to AUDIT-C (Alcohol Use Disorders Identification Test–Consumption), GAD-7 (Generalized Anxiety Disorder-7), and PHQ-9 (Patient Health Questionnaire-9). Columns indicate FreeSurfer-derived brain regions. Colors denote standardized association strength (z-scores), with blue indicating negative associations and red indicating positive associations. Z-scoring was applied across all regions to emphasize relative patterns.

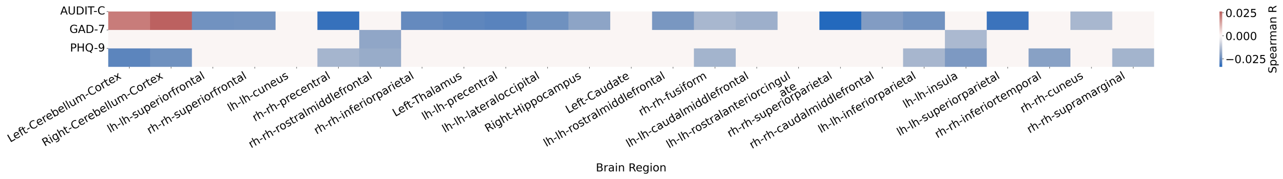


Figure 13: Top 25 brain regions ranked by absolute effect size of Spearman's $\rho$ correlations between FreeSurfer-derived structural features and symptom severity scores. Rows correspond to AUDIT-C (Alcohol Use Disorders Identification Test–Consumption), GAD-7 (Generalized Anxiety Disorder-7), and PHQ-9 (Patient Health Questionnaire-9). Columns indicate individual FreeSurfer brain regions. Colors represent the strength and direction of correlations (blue = negative, red = positive), with intensity reflecting absolute correlation magnitude.

Table 14: ElasticNet-derived explained variance for significant embedding dimensions.

| Score | Dimension | R2 |
|---|---|---|
| AUDIT-C sum score | 224 | 0.110 |
| AUDIT-C sum score | 162 | 0.395 |
| AUDIT-C sum score | 204 | 0.227 |
| AUDIT-C sum score | 155 | 0.280 |
| AUDIT-C sum score | 86 | 0.224 |
| AUDIT-C sum score | 73 | 0.131 |
| AUDIT-C sum score | 62 | 0.097 |
| AUDIT-C sum score | 87 | 0.229 |
| AUDIT-C sum score | 122 | 0.358 |
| AUDIT-C sum score | 216 | 0.194 |
| AUDIT-C sum score | 182 | 0.172 |
| AUDIT-C sum score | 102 | 0.328 |
| AUDIT-C sum score | 112 | 0.229 |
| AUDIT-C sum score | 223 | 0.200 |
| AUDIT-C sum score | 31 | 0.182 |
| AUDIT-C sum score | 177 | 0.290 |
| AUDIT-C sum score | 61 | 0.098 |
| AUDIT-C sum score | 173 | 0.108 |
| GAD-7 sum score | 100 | 0.098 |
| GAD-7 sum score | 241 | 0.378 |
| GAD-7 sum score | 217 | 0.215 |
| GAD-7 sum score | 58 | 0.281 |
| PHQ-9 sum score | 76 | 0.104 |
| PHQ-9 sum score | 217 | 0.215 |
| PHQ-9 sum score | 246 | 0.204 |
| PHQ-9 sum score | 100 | 0.098 |
| PHQ-9 sum score | 205 | 0.401 |
| PHQ-9 sum score | 132 | 0.267 |
| PHQ-9 sum score | 225 | 0.105 |
| PHQ-9 sum score | 231 | 0.336 |
| PHQ-9 sum score | 6 | 0.220 |
| PHQ-9 sum score | 233 | 0.262 |
| PHQ-9 sum score | 45 | 0.214 |
| PHQ-9 sum score | 91 | 0.223 |
| PHQ-9 sum score | 227 | 0.383 |
| PHQ-9 sum score | 216 | 0.194 |

##### 3.7 Classification Baseline

| Group | Confounders only | | Freesurfer only | |
|---|---|---|---|---|
| | **BACC** | **AUC** | **BACC** | **AUC** |
| PHQ-9 (1) | 58 ± 1 | 61 ± 1 | 58 ± 1 | 61 ± 1 |
| PHQ-9 (2) | 61 ± 2 | 65 ± 2 | 61 ± 3 | 65 ± 2 |
| PHQ-9 (3) | 57 ± 9 | 59 ± 15 | 65 ± 3 | 70 ± 3 |
| PHQ-9 (4) | 57 ± 16 | 59 ± 29 | 69 ± 7 | 76 ± 7 |
| MDD ICD-10 | 57 ± 3 | 61 ± 3 | 58 ± 3 | 61 ± 3 |
| MDD (diag.) | 58 ± 1 | 61 ± 1 | 58 ± 1 | 61 ± 2 |
| GAD-7 (2) | 58 ± 2 | 61 ± 2 | 58 ± 2 | 61 ± 2 |
| GAD-7 (3) | 57 ± 5 | 61 ± 7 | 57 ± 4 | 61 ± 4 |
| GAD-7 (4) | 62 ± 7 | 66 ± 13 | 63 ± 4 | 69 ± 5 |
| ANX ICD-10 | 56 ± 4 | 60 ± 4 | 57 ± 3 | 60 ± 3 |
| ANX (diag.) | 56 ± 2 | 58 ± 2 | 56 ± 2 | 58 ± 2 |
| AUDIT-C (8) | 60 ± 3 | 65 ± 3 | 60 ± 3 | 63 ± 3 |
| AUDIT-C (9) | 63 ± 3 | 68 ± 4 | 62 ± 4 | 66 ± 3 |
| AUDIT-C (≥ 10) | 63 ± 3 | 68 ± 3 | 64 ± 3 | 69 ± 3 |
| AUD ICD-10 | 58 ± 8 | 57 ± 11 | 58 ± 5 | 64 ± 6 |

Table 15: Balanced accuracy (BACC; computed at a decision threshold of 0.5) and area under the receiver operating characteristic curve (AUC) for classification across all diagnostic outcomes in the UKB test set. Models included: *Confounders only* (sex, age, age-squared, and age–sex interaction), *FreeSurfer only* (Freesurfer-derived brain features with covariates). Values represent mean ± standard deviation across 10-fold stratified cross-validation.

##### 3.8 Classification Performance Details

Table 16: Balanced accuracy (BACC; computed at a decision threshold of 0.5) distributions across 10-fold stratified cross-validation for all diagnostic outcomes. Columns show the minimum, maximum, mean, median, and standard deviation of BACC values across folds. Model configurations include: *Confounders only* (sex, age, age-squared, age–sex interaction), *Deviation 256d + covariates* (256-dimensional brain deviations with covariates).

| Target | Config | Min | Max | Mean | Median | Std |
|---|---|---|---|---|---|---|
| MDD ICD-10 | Confounders only | 0.53 | 0.65 | 0.57 | 0.57 | 0.03 |
| MDD ICD-10 | Deviation 256d + covariates | 0.58 | 0.67 | 0.62 | 0.62 | 0.03 |
| PHQ-9 (1) | Confounders only | 0.56 | 0.60 | 0.58 | 0.58 | 0.01 |
| PHQ-9 (1) | Deviation 256d + covariates | 0.56 | 0.61 | 0.59 | 0.59 | 0.02 |
| PHQ-9 (2) | Confounders only | 0.58 | 0.64 | 0.61 | 0.61 | 0.02 |
| PHQ-9 (2) | Deviation 256d + covariates | 0.60 | 0.67 | 0.63 | 0.62 | 0.02 |
| PHQ-9 (3) | Confounders only | 0.47 | 0.73 | 0.62 | 0.62 | 0.07 |
| PHQ-9 (3) | Deviation 256d + covariates | 0.51 | 0.68 | 0.61 | 0.61 | 0.06 |

*Continued on next page*

| Target | Config | Min | Max | Mean | Median | Std |
|---|---|---|---|---|---|---|
| PHQ (4) | Confounders only | 0.31 | 0.77 | 0.59 | 0.66 | 0.18 |
| PHQ (4) | Deviation 256d + covariates | 0.56 | 0.81 | 0.70 | 0.71 | 0.07 |
| GAD (2) | Confounders only | 0.55 | 0.60 | 0.58 | 0.59 | 0.02 |
| GAD (2) | Deviation 256d + covariates | 0.55 | 0.61 | 0.58 | 0.59 | 0.02 |
| GAD (3) | Confounders only | 0.53 | 0.62 | 0.58 | 0.57 | 0.03 |
| GAD (3) | Deviation 256d + covariates | 0.58 | 0.65 | 0.61 | 0.60 | 0.02 |
| GAD (4) | Confounders only | 0.53 | 0.70 | 0.65 | 0.67 | 0.05 |
| GAD (4) | Deviation 256d + covariates | 0.53 | 0.71 | 0.62 | 0.63 | 0.05 |
| ANX ICD-10 | Confounders only | 0.50 | 0.60 | 0.56 | 0.56 | 0.03 |
| ANX ICD-10 | Deviation 256d + covariates | 0.55 | 0.62 | 0.58 | 0.58 | 0.02 |
| AUD ICD-10 | Confounders only | 0.49 | 0.65 | 0.57 | 0.56 | 0.06 |
| AUD ICD-10 | Deviation 256d + covariates | 0.55 | 0.68 | 0.62 | 0.62 | 0.04 |
| AUDIT-C (8) | Confounders only | 0.56 | 0.64 | 0.60 | 0.59 | 0.03 |
| AUDIT-C (8) | Deviation 256d + covariates | 0.53 | 0.64 | 0.60 | 0.61 | 0.03 |
| AUDIT-C (9) | Confounders only | 0.58 | 0.68 | 0.63 | 0.64 | 0.04 |
| AUDIT-C (9) | Deviation 256d + covariates | 0.57 | 0.69 | 0.64 | 0.64 | 0.04 |
| AUDIT-C (10) | Confounders only | 0.57 | 0.68 | 0.63 | 0.63 | 0.03 |
| AUDIT-C (10) | Deviation 256d + covariates | 0.60 | 0.69 | 0.66 | 0.66 | 0.03 |
| MDD (diag.) | Confounders only | 0.56 | 0.60 | 0.58 | 0.58 | 0.01 |
| MDD (diag.) | Deviation 256d + covariates | 0.56 | 0.61 | 0.59 | 0.59 | 0.01 |
| ANX (diag.) | Confounders only | 0.53 | 0.59 | 0.56 | 0.56 | 0.02 |
| ANX (diag.) | Deviation 256d + covariates | 0.54 | 0.59 | 0.57 | 0.57 | 0.02 |

Table 17: Area under receiver operating characteristic curve (AUC) distributions across 10-fold stratified cross-validation for all diagnostic outcomes. Columns show the minimum, maximum, mean, median, and standard deviation of balanced accuracy (BACC; computed at a decision threshold of 0.5) values across folds. Model configurations include: *Confounders only* (sex, age, age-squared, age–sex interaction), *PRS + covariates* (polygenic risk scores with covariates), *Deviation 256d + covariates* (256-dimensional brain deviations with covariates), and *PRS + Deviation 256d + covariates* (combined model).

| Target | Config | Min | Max | Mean | Median | Std |
|---|---|---|---|---|---|---|
| MDD ICD-10 | Confounders only | 0.53 | 0.68 | 0.61 | 0.61 | 0.04 |
| MDD ICD-10 | Deviation 256d + covariates | 0.62 | 0.72 | 0.66 | 0.66 | 0.03 |
| PHQ (1) | Confounders only | 0.58 | 0.62 | 0.61 | 0.61 | 0.01 |
| PHQ (1) | Deviation 256d + covariates | 0.59 | 0.64 | 0.61 | 0.61 | 0.01 |
| PHQ (2) | Confounders only | 0.62 | 0.71 | 0.65 | 0.65 | 0.02 |
| PHQ (2) | Deviation 256d + covariates | 0.63 | 0.70 | 0.67 | 0.67 | 0.02 |
| PHQ (3) | Confounders only | 0.48 | 0.75 | 0.68 | 0.69 | 0.08 |
| PHQ (3) | Deviation 256d + covariates | 0.55 | 0.74 | 0.66 | 0.68 | 0.07 |
| PHQ (4) | Confounders only | 0.13 | 0.89 | 0.59 | 0.71 | 0.30 |

*Continued on next page*

| Target | Config | Min | Max | Mean | Median | Std |
|---|---|---|---|---|---|---|
| PHQ (4) | Deviation 256d + covariates | 0.64 | 0.86 | 0.75 | 0.74 | 0.08 |
| GAD (2) | Confounders only | 0.58 | 0.64 | 0.61 | 0.62 | 0.02 |
| GAD (2) | Deviation 256d + covariates | 0.59 | 0.64 | 0.61 | 0.62 | 0.02 |
| GAD (3) | Confounders only | 0.57 | 0.67 | 0.63 | 0.63 | 0.03 |
| GAD (3) | Deviation 256d + covariates | 0.58 | 0.69 | 0.63 | 0.63 | 0.03 |
| GAD (4) | Confounders only | 0.55 | 0.77 | 0.70 | 0.73 | 0.07 |
| GAD (4) | Deviation 256d + covariates | 0.54 | 0.73 | 0.65 | 0.65 | 0.06 |
| ANX ICD-10 | Confounders only | 0.52 | 0.65 | 0.59 | 0.59 | 0.05 |
| ANX ICD-10 | Deviation 256d + covariates | 0.58 | 0.68 | 0.63 | 0.63 | 0.04 |
| AUD ICD-10 | Confounders only | 0.46 | 0.68 | 0.59 | 0.62 | 0.08 |
| AUD ICD-10 | Deviation 256d + covariates | 0.52 | 0.70 | 0.65 | 0.66 | 0.05 |
| AUDIT-C (8) | Confounders only | 0.58 | 0.70 | 0.65 | 0.64 | 0.04 |
| AUDIT-C (8) | Deviation 256d + covariates | 0.57 | 0.70 | 0.65 | 0.64 | 0.04 |
| AUDIT-C (9) | Confounders only | 0.61 | 0.75 | 0.68 | 0.69 | 0.04 |
| AUDIT-C (9) | Deviation 256d + covariates | 0.63 | 0.76 | 0.69 | 0.69 | 0.04 |
| AUDIT-C (10) | Confounders only | 0.62 | 0.71 | 0.68 | 0.69 | 0.03 |
| AUDIT-C (10) | Deviation 256d + covariates | 0.65 | 0.75 | 0.72 | 0.72 | 0.03 |
| MDD (diag.) | Confounders only | 0.58 | 0.63 | 0.61 | 0.61 | 0.02 |
| MDD (diag.) | Deviation 256d + covariates | 0.60 | 0.64 | 0.62 | 0.62 | 0.01 |
| ANX (diag.) | Confounders only | 0.55 | 0.62 | 0.58 | 0.58 | 0.02 |
| ANX (diag.) | Deviation 256d + covariates | 0.55 | 0.62 | 0.60 | 0.60 | 0.02 |

[3] Junnan Li, Ramprasaath R. Selvaraju, Akhilesh D. Gotmare, Shafiq Joty, Caiming Xiong, and Steven C.H. Hoi. Align before fuse: vision and language representation learning with momentum distillation. In *Proceedings of the 35th International Conference on Neural Information Processing Systems*, NIPS '21, Red Hook, NY, USA, 2021. Curran Associates Inc. ISBN 9781713845393.

[4] Jielin Qiu, Jiacheng Zhu, Shiqi Liu, William Han, Jingqi Zhang, Chaojing Duan, Michael A. Rosenberg, Emerson Liu, Douglas Weber, and Ding Zhao. Automated cardiovascular record retrieval by multimodal learning between electrocardiogram and clinical report. In Stefan Hegselmann, Antonio Parziale, Divya Shanmugam, Shengpu Tang, Mercy Nyamewaa Asiedu, Serina Chang, Tom Hartvigsen, and Harvineet Singh, editors, *Proceedings of the 3rd Machine Learning for Health Symposium*, volume 225 of *Proceedings of Machine Learning Research*, pages 480–497. PMLR, 10 Dec 2023. URL `https://proceedings.mlr.press/v225/qiu23a.html`.

[5] Fernando Pérez-García, Rachel Sparks, and Sébastien Ourselin. TorchIO: A python library for efficient loading, preprocessing, augmentation and patch-based sampling of medical images in deep learning. *Computer Methods and Programs in Biomedicine*, 208:106236, 2021. doi: 10.1016/j.cmpb.2021.106236.

[6] Adam Paszke, Sam Gross, Soumith Chintala, Gregory Chanan, Edward Yang, Zachary DeVito, Zeming Lin, Alban Desmaison, Luca Antiga, and Adam Lerer. Automatic differentiation in PyTorch. 2017.

[7] Bruce Fischl. Freesurfer. *NeuroImage*, 62(2):774–781, 2012. ISSN 1053-8119. doi: 10.1016/j.neuroimage.2012.01.021. 20 YEARS OF fMRI.

[8] B.W. Silverman. *Density Estimation for Statistics and Data Analysis.* 02 2018. ISBN 9781315140919. doi: 10.1201/9781315140919.

[9] E. WEISZFELD. Sur le point pour lequel la somme des distances de n points donnés est minimum. *Tohoku Mathematical Journal, First Series*, 43:355–386, 1937.

[10] John D. Storey. A direct approach to false discovery rates. *Journal of the Royal Statistical Society Series B: Statistical Methodology*, 64(3):479–498, 2002. doi: 10.1111/1467-9868.00346.
